# The use of data and analytics for health sector priority-setting in Ghana: a qualitative study

**DOI:** 10.1101/2025.07.17.25331378

**Authors:** Emmanuelle A. Dankwa, Catherine Wambura, Marwatunnisa Al Mubarokah, Christina J. Matta, Bellama Gado, Joyce Komesuor, John Van Savage, Anna Makido, Frank Baiden, Abdisalan M. Noor

## Abstract

Over the last few decades, Ghana has implemented health sector reforms to improve health care access in alignment with Sustainable Development Goal 3. However, Ghana’s health sector continues to face challenges alongside mounting healthcare needs and persistent overspending of the healthcare budget. These challenges are exacerbated by recent cuts in external aid to Ghana, illustrating the importance of effective health sector priority-setting (HSPS). HSPS describes the process of determining how to best allocate resources to maximize population health outcomes among various alternative, competing health needs and interventions. Although Ghana has a history of priority-setting initiatives, there is no authoritative source on the current landscape of HSPS in Ghana. This study sought to fill this gap by offering a detailed analysis of current HSPS processes at the national level in Ghana, focusing on the use of data and analytics in informing HSPS. The study uses data from key informant interviews and a literature review to provide information on the processes and institutions involved in HSPS in Ghana, data and analytics used in HSPS, and successes, challenges, and recommendations.

Findings show several good aspects of HSPS in Ghana, including demonstrated political commitment and attempts at a bottom-up approach. However, challenges remain, including political and development partner influences, lack of a nationally owned, unified national health information system (HIS) infrastructure, and a lack of capacity and demand for advanced analytics in decision-making. Recommendations include developing a sustainable, integrated HIS that is nationally owned, developing data and analytic capacity within the health sector, improving participation of communities and subnational governments in priority-setting, facilitating closer collaboration between wider sectoral and program-specific priority-setting, and committing to a long-term domestic financing plan for healthcare.

## Introduction

Ghana has passed policies and reforms to improve healthcare access and establish a health system based on the Universal Health Coverage (UHC) principles. Since the 1990s, Ghana has implemented reforms within its health sector, including adopting decentralized governance and collaborative frameworks that engage various levels of government, civil society organizations (CSOs), the private sector, and international organizations. These reforms aimed to enhance service delivery at the community level while addressing the health needs across Ghana’s urban and rural areas [1,2].

Ghana allocates 2.02% of its GDP to healthcare and this is channeled mainly through its Ministry of Health (MoH) [3]. The health funding structure relies on multiple sources, with the government of Ghana being the primary contributor at 54% of the budget in 2023 and development partner funding contributing 20% of the budget in that year [3,4]. A key challenge of healthcare spending in Ghana is the overspending of the healthcare budget in recent years: between 2019 and 2022, the MoH expenditure exceeded the budget by 6%-13% [3], demonstrating that there are more healthcare expenses than allowed by the budget. A related challenge is the underfunding of the budget, resulting in important health needs being unmet.

Given these challenges and with the recent cuts in external aid to Ghana [5], health sector priority-setting (HSPS) is critical for the Ghanaian context. HSPS is the process of determining how to efficiently and effectively allocate limited resources to meet population health needs. It is the third stage of a country’s health policy and planning processes which typically consists of eight stages: population consultation, health situational analysis, HSPS, national health strategic planning, operational planning, costing, budgeting, and monitoring and evaluation [6].

Ghana has a history of priority-setting initiatives, including the International Decision Support Initiative (iDSI) [7] and the recent launch of the first strategy for Health Technology Assessment [8,9]. Despite these emerging initiatives and the relevance of HSPS for Ghana’s context, there is no authoritative source on the current landscape of HSPS in Ghana. References on the subject either focus on a single HSPS initiative [e.g., 7,9,10] or priority-setting for a single aspect of the health sector [e.g., 11] No source, as far as we know, explains the HSPS process at the national level, and how data is used to inform the processes. This presents a gap in understanding of data use for HSPS in Ghana, making it challenging to propose any recommendations for improvement.

This paper sought to address this gap by offering a detailed analysis of current HSPS processes at the national level in Ghana, focusing on the use of data and analytics in HSPS. It combines an extensive literature review with key informant interviews (KIIs) to provide information on the following in the context of HSPS: Ghana’s health governance structure, processes, relevant organizations, data and analytics, successes, challenges and recommendations. The paper is outlined as follows: 1) The methods are discussed. 2) Then, to establish context, the health governance and health delivery system of Ghana and priority-setting processes are discussed based on the literature review. 3) Results from the interviews are then presented, organized by four themes: institutions involved in Ghana’s HSPS; priority-setting processes; use of data and analytics in HSPS; and successes and challenges of HSPS. 4) Recommendations for improvement of HSPS are also suggested, based on the literature review, interviews and evidence from other countries. 5) Finally, a discussion summarizing key points of the paper is presented.

## Methods

The study used data from two complementing sources: a literature review and KIIs. The KIIs were intended to fill in any information gaps left by the extensive literature review.

### Literature Review

The literature review included peer-reviewed research articles, grey literature, and essential policy documents to explore Ghana’s HSPS processes. This desk review examined crucial aspects, such as HSPS implementation steps, the roles of involved individuals and institutions, data and analytics usage, and identified challenges, successes, and potential improvements. The search for relevant sources covered prominent public health databases, including Elsevier, PubMed, and Google Scholar, focusing on documents published within the past decade to ensure up-to-date insights. Additional information was obtained from the MoH and Ghana Health Service (GHS) websites focusing on health governance and how the different agencies and departments are involved in the HSPS processes.

### Qualitative Study

#### Study Design

The qualitative study involved KIIs and used a semi-structured interview approach with a combination of purposive and snowball sampling of participants. KIIs were conducted in August 2024 in Accra, Ghana with individuals directly or indirectly involved in HSPS to gather contemporary insights on HSPS practices in Ghana. Interviews were guided by questions about context, key actors, processes, successes, and challenges. Interview guides are available as a supplementary material in S2 Text. Questions were structured to fill the gaps identified in the literature review. The style and content of questions were adapted to suit the background, expertise, and institution of participants. The semi-structured approach allowed for the flexibility for follow-up questions to explore any relevant themes which arose during interviews if not captured in the prepared questions.

#### Study Population and Participant Recruitment

Eight participants were selected through a combination of purposive sampling and snowball sampling, from governmental institutions, academia, and partner organizations. This enabled a targeted selection of participants who are well-versed in the health sector in Ghana, most of whom had worked within their organizations and roles at senior levels, allowing for participants to paint a robust picture of Ghana’s priority-setting processes. Participants were current or former employees of two governmental agencies (MoH and GHS), two development partner or donor organizations, (World Health Organization (WHO) and Program for Appropriate Technology in Health (PATH)) and one academic institution experienced in collaborating with the GHS (University of Health and Allied Sciences Ghana). Additional participants were not sought after saturation had been achieved by the last interview [12].

#### Data Collection

Interviews were conducted separately for each participant by two to four interviewers. Interviews were in English and were conducted in-person or online using Zoom, according to participants’ preferences. Interviews were recorded through notetaking by two researchers and were summarized by one researcher who compared notes to ensure consistency. Summary involved paraphrasing participants’ responses while retaining relevant details. To ensure accuracy, prior to being analyzed, the summarized notes were again reviewed by one researcher who was present at all interviews.

#### Data Analysis

The analysis was conducted collaboratively using a thematic approach. The process began with an initial familiarization process, during which a subset of summarized interview notes was read with purposeful overlap to facilitate discussion. An initial codebook was developed based on the preliminary summaries and insights, which was later updated accordingly. Each interview was then independently coded by two researchers, followed by a comparison of codes and discussions to resolve any discrepancies. NVivo 14 software was used to explore and organize the data, enabling the examination of patterns both within individual codes and across the entire dataset. To enhance rigor, additional examination of themes and codes was conducted by two other researchers on the team to ensure inter-coder reliability for consistent coding. This iterative, collaborative analysis process allowed each team member to analyze specific codes and themes, with analysis interpretations regularly discussed within the research team to refine interpretations.

#### Reflexivity Statement

The research was conducted collaboratively using a reflexive approach, with a diverse team of various races, ethnicities, and nationalities, who brought varying backgrounds, expertise, and personal experiences. This provided a balanced foundation for analyzing complex processes and allowed for an examination of the priority-setting processes through multiple perspectives, adding depth and breadth to our findings. The stakeholder consultations were conducted in English, the primary language in Ghana, facilitating effective communication with participants. KIIs were led by researchers and native Ghanaians at the University of Health and Allied Sciences which helped capture subtle nuances and foster trust with key informants. Analysis was led by researchers at the Harvard T.H. Chan School of Public Health and validated by University of Health and Allied Sciences colleagues. The team’s collective expertise in literature review, qualitative methodologies, and health systems strengthened the study’s rigor and systematic approach. Despite efforts to ensure objectivity, the researchers are aware that inherent biases may subtly influence the interpretation of the findings. Transparency, therefore, is prioritized in presenting conclusions, with recognition that the research team’s backgrounds may shape perspectives and analysis.

#### Ethical Review

Ethical approval for this study was not required by the Institutional Review Board of the Harvard T.H. Chan School of Public Health due to the determination that the study did not constitute human subjects research as defined by Department of Health and Human Services regulations or Food and Drug Administration regulations. Participants provided written informed consent to participate.

### Information from the Literature

#### Health Governance Structure

The MoH is the primary policymaking and regulatory body responsible for central-level activities, including policymaking, regulation, and planning coordination [13]. Authority from the Ministry of Health is delegated to its agencies, including the Ghana Health Service (GHS), Teaching Hospitals, and faith-based service providers. The GHS is the main implementation arm on government healthcare policies and administrates healthcare delivery, controlling the operations of most government health care facilities. The GHS comprises 11 divisions, including the Public Health Division, the Research and Development Division (RDD) and the Policy, Planning, Monitoring, and Evaluation Division (PPME) (Fig. A in S1 Text). Each division is headed by a divisional director who reports directly to the Director General. The Public Health Division oversees disease-specific control programs, such as the Tuberculosis (TB) Control Program, which are headed by program managers.

#### Health Delivery System

The health delivery system is organized into three tiers (Fig. B in S1 Text). The primary level includes district hospitals, sub-district health centers, and community-based health planning services [13]. This level aims to provide community members with basic essential primary health care services, which are critical for the attainment of UHC. The secondary level, represented by regional hospitals, offers public health and clinical services and acts as referral centers for primary level facilities. At the tertiary level, specialized care delivery is provided through teaching, university, and psychiatry hospitals which serve as hubs for training health professionals and provide services to clients referred from the other two levels [13]. At the primary and secondary levels, health directorates manage health service delivery [14].

The private sector and CSOs also support the government’s efforts in healthcare service delivery, accounting for 19% of outpatient department visits, with facilities predominantly located in urban and peri-urban areas [13]. Faith-based organizations also support governmental efforts to provide primary and secondary level care by establishing and operating health facilities [13].

#### Health Service Data

Ghana has several health data sources, such as the Demographic and Health Survey (DHS) data, Global Burden of Disease, National Health Insurance Scheme data, Lightwave Health Information Management System (LHIMS), and the District Health Information Management System (DHIMS2) which is the main repository of health service data for the GHS and MoH [9,15]. All health sector data in Ghana is managed by the Centre for Health Information Management under the Information Monitoring and Evaluation Department of the GHS’ PPME [9]. However, there is no composite database for all healthcare data and existing systems have limited interoperability; therefore, data remains largely compartmentalized [9].

For details on the structure, function and challenges associated with major health data sources in Ghana, see the supplementary material in S1 Text.

### Priority-Setting Processes

#### Overview

Ghana’s priority-setting process is rooted in a constitutional mandate spearheaded by the National Development Planning Commission, which guides public and civil society organizations in shaping strategic plans [13]. The Health Sector Medium-Term Development Plan (HSMTDP) is the blueprint for Ghana’s health system, guiding investments, interventions and reforms.

The 2022-2025 HSMTDP outlines the health sector’s priorities for the period, which were determined based on the following [13]: 1) a review of the previous (2018-2021) HSTMDP, 2) a holistic assessment of the health sector conducted in 2020 [16], and 3) the Aide Memoire, a document summarizing priority decisions made by government and development partners at the annual health sector summit. The development of the 2022-2025 HSMTDP was also guided by the Ghana National Health Policy [2] and the UHC Roadmap for Ghana (2020-2030). An annual Program of Work (PoW) document outlines the action plan for implementation of the HSMTDP for each year. For example, the 2024 PoW [17] outlines priority areas for focus in 2024, aligning with the 2022-2025 HSMTDP.

#### Health System Assessment

The MoH conducts a performance assessment of interventions and health activities implemented according to the previous strategic plan and uses the assessments inform priorities for a new plan. In addition to a national assessment of the health sector (known as the Holistic Assessment), area-specific assessments, such as on public expenditure and public health emergencies, are conducted [18]. For example, a review of the 2018-2021 HSMTDP informed priorities for the 2022-2025 HSMTDP. The MoH collaborates with its agencies, development partners and other health stakeholders in the development of these priorities [13].

#### The Health Summit

Findings on the assessments of the implementation of the previous health strategic plan are presented to health sector stakeholders at an annual health sector planning meeting known as the annual health summit, hosted by the MoH [18,19]. For example, the 2024 summit presented results from the mid-term review of the HSMTDP, providing an opportunity for the MoH, GHS, and other invited health sector stakeholders to discuss the report and share insights into the health system’s performance and effectiveness based on stated policy objectives outlined in the HSMTDP and the UHC Roadmap of 2020 – 2030 [20]. Within the health summit, a “business meeting” is held to finalize priorities. This meeting is held between the management of the MoH, directors of MoH agencies, and development partners [20]. Priorities finalized in this meeting are summarized into the *aide memoire*, which informs the PoW for the upcoming year [8].

#### Inter-Agency Leadership Committee

Another important avenue for HSPS is a regular meeting between key health sector leaders in government, known as the Inter-Agency Leadership Committee. It exists to facilitate unity in the health sector by providing a platform for the exchange of ideas which will “influence policy and provide overall strategic direction in the health sector” [18]. It meets at least quarterly and comprises the Ministers of Health, Chief Director of the MoH, heads of MoH agencies, Director General of the Ghana Aids Commission (the Commission is under the Office of the President and is not an agency of the MoH) and the PPME Director, who is the committee’s secretary. It is chaired by the Minister of Health. Other stakeholders external to the government may be invited to the Committee’s meetings as needed.

#### Health Sector Working Group

The Health Sector Working Group is a forum for engagement between government health sector leaders and sector partners external to the government, including civil society organizations, local and international non-governmental organizations, development partners and the private sector [18]. The Working Group exists to facilitate policy dialogues among key sector partners, monitor programs such that they achieve desired aims, improve the alignment of development partners’ activities with government priorities, review resource allocation and discuss and agree on the annual PoW [18]. These meetings are held quarterly and chaired by the Minster of Health.

### Gaps in the Literature

Regarding the health system assessments which precede and influence priority-setting, the literature did not have information on how exactly these assessments are conducted, the data sources and analytics used to perform these assessments, the agencies which provide data for these assessments, and how district or regional level assessments inform national assessments. The literature does not discuss successes or challenges associated with HSPS in Ghana. Consequently, there are no recommendations for the improvement of HSPS in Ghana. The qualitative analysis presented in the next section was intended to address these gaps.

## Results from Qualitative Analysis

Four main themes were identified from the KIIs: 1) institutions involved in HSPS in Ghana, 2) HSPS processes, 3) data and analytics for HSPS and 4) successes and challenges in HSPS. These are now discussed by theme (Table 1). Interview notes are available in S3 Text.

**Table 1.** Themes identified from qualitative key information interviews.

| Themes | Sub-themes |
| --- | --- |
| Institutions involved in health sector priority-setting | • Governmental organizations and agencies<br>• Development partners |
| Priority-setting processes | • Health system assessments<br>• Processes between realization of assessments and formation of priorities |
| Data and Analytics | • Data analysis processes<br>• Data and analytic capacity |
| Successes and Challenges | • Setting and implementing priorities<br>• Data and analytics |

### Theme 1: Institutions Involved in Ghana’s HSPS

#### Literature gap

It is not clear from the literature how development partners engage with HSPS outside the health summit.

#### What this study adds

The MoH engages with development partners including the Global fund, World Bank, United Nations International Children’s Emergency Fund (UNICEF), Korea International Cooperation Agency [Interviews 2, 3], Japan International Cooperation Agency [Interview 3], WHO [Interviews 2, 3, 4, 7], USAID (Interview 4) and PATH [Interview 1] at various stages in the HSPS process. It is notable that USAID has been effectively dissolved since March, 2025 [5]; however, its role in Ghana’s priority-setting processes prior to this date is explained. Development partners support the MoH technically and financially to assess the health system and identify critical gaps, plan and implement interventions to fill these gaps, evaluate implementations [Interviews 1, 7], manage data, collect data, and develop staff capacity in data analytics [Interview 2]. For example, the WHO has supported the country in generating relevant data for conducting a SCORE assessment—a tool to help countries strengthen data systems and monitor progress toward health goals [Interview 7]. PATH also conducts landscaping to identify gaps in the health sector and supports the MoH to address the issue within PATH’s capacities [Interview 1]. The WHO and PATH support the MoH in its implementation of the priorities set during the health summit [Interviews 1, 7]. USAID provided technical assistance in generating, analyzing, and utilizing data during priority-setting processes [Interview 4].

GHS also works with academic institutions to collect and analyze data with outputs informing some GHS research reports to the MoH. For example, the University of Ghana School of Public Health, University of Health and Allied Sciences, and Kwame Nkrumah University of Science and Technology work with the GHS as researchers and consultants to generate and disseminate data [Interviews 2, 6].

### Theme 2: Priority-Setting Processes

#### Health System Assessments and Reviews

##### Literature gap

The literature does not explain how assessments are conducted within the GHS and other MoH agencies; in particular, the coordination between GHS divisions, across

GHS governance levels (district, regional, national) and between the GHS and development partners.

##### What this study adds

To prepare the assessment report by the GHS, each GHS division and program at the national level conducts an annual performance review (APR) of its activities and prepares an assessment report [Interviews 3, 5]. These APRs assess performance indicators against priorities [Interview 5] as outlined in the PoW for the previous year [Interview 2]. Data for these assessments are mainly sourced from DHIMS2 [Interview 4], though demographic data are also used [Interview 2]. Divisional directors present and discuss their reports in a meeting chaired by the Director General of the GHS [Interview 3]. At the district and regional levels of health service delivery, health directorates also conduct thorough APRs [Interview 5], in addition to quarterly activity assessments [9]. District assessments are collated at the regional level and regional assessments are collated at the national level [Interview 5].

Aside these assessments, as a monitoring strategy, the PPME also conducts a quarterly assessment of health services based on DHIMS2 data [Interview 5]. These assessments are collated by the PPME into an overall GHS assessment [Interviews 3, 5]. Therefore, the national GHS assessment is a composite of divisional and sub-national assessments.

### From Assessments to Priorities

#### Literature gap

The literature is not clear on the processes which occur between the final assessment and the final set of priorities.

#### What this study adds

The process by which assessments influence priorities was explained as follows. The GHS Director General decides which aspects of the GHS assessment to highlight at a meeting chaired by the Minister of Health and attended by other MOH agency heads (e.g., heads of teaching hospitals) who also report on their agency assessments [Interview 3]. Decisions by the Director General of the GHS on what to highlight at the MoH meeting are generally based on the burden of the health problem as showed by the assessments [Interview 3] or on emerging health needs [Interview 5].

After assessments from the GHS and other MoH agencies are presented to the Minister of Health, an ad hoc committee led by the PPME director is formed and its output is an assessment of the health situation, which contributes to the *aide memoire* [Interview 4]. The constitution of the committee depends on the health indicators or issues that were highlighted for attention, based on presentations by agencies [Interview 4]. The Minister of Health presents this assessment to government leaders [Interview 3]. During the annual health summit, all agencies of the MoH, including the GHS, are expected to report on the assessments of their PoW for the previous year [Interview 2].

Attendance to the last day of the summit, referred to as thelJbusiness meeting, is limited to the minister, relevant agencies, select staff from the GHS and the MoH at the national level and development partners, including PATH [Interviews 1, 5]. This meeting serves as a deliberative process to assign scores to various priorities, which ultimately determines a set of priorities based on the highest score across all indicators [Interviews 1, 7]. The *aide memoir* outlines the agreed priorities and funding commitments between the government and partners [Interview 4, 5] for the subsequent year. Some development partners such as PATH do not directly influence priority decisions at the health summit but rather do so at the regional and national levels [Interview 1].

Several factors shape the determination of priorities. These include the seriousness of the health problem assessed by the associated burden of disease; resource or funding availability; emerging health trends; and the global health agenda, particularly the Sustainable Development Goal 3 and the UHC roadmap [Interviews 1, 4, 5, 8]. The latter factor is because Ghana is keen to remain connected to the global agenda and related international programs [Interview 2]. In terms of availability of resources, for instance, out of the funds allocated to the health sector, close to 70% goes to personal emoluments, leaving only 30% for activities within the ministry, which is insufficient [Interview 2].

Additionally, donors usually have their own priorities; for example, USAID prioritized health system strengthening, monitoring, and evaluation, while the World Bank prioritizes programs such as HIV and TB [Interviews 2, 5]. PATH also has priority focus areas but regularly conducts its own situational analysis to identify gaps and uses this information to determine priority areas requiring its support [Interview 1]. As an additional example, an interviewee noted that in the past, one funder only supported work in five (out of ten) regions because the funder had designated those as its priority regions, due to poverty levels there [Interview 5].

Given that priorities-related activities cannot be conducted without adequate financial support, and local funding is severely limited, Ghana is heavily reliant on donors for priority-setting and implementation [Interviews 3, 5]. Donor influences, in this respect, weaken the MoH’s ability to ensure data-driven health priority-setting in alignment with country needs [Interviews 2, 3, 5]. However, if donor funding may need to be repurposed due to unforeseen circumstances, the Ministry of Finance could reprioritize where to use the funds and inform the development partner about the change in the purpose of the funds [Interview 5]. For example, WHO funds for the Maternal and Child Health and Nutrition Project were repurposed to fund COVID-19 control efforts [Interview 5].

Political influences on the priority-setting process further exacerbate this issue of misalignment of priorities and country needs [Interviews 1, 5]. Health priorities have sometimes been revised to incorporate manifesto promises, even if that did not reflect prevailing needs. For example, during the 2018 annual health summit, the Vice President mentioned the use of drones to deliver vaccines which was not a health sector priority then, but policies were later developed to accommodate the Vice President’s comments [Interview 5].

### Summary of HSPS Processes

The step-by-step process of priority setting at the national level in Ghana, as explained by the key informants and from the literature, is summarized in Fig. 1. The figure accounts for the events, processes, players, data used and products at each stage of HSPS in Ghana at the national level.

**Fig. 1.** Ghana’s step-by-step priority-setting processes.

### Theme 3: Data and Analytics Used in Priority-Setting

#### Data Analysis Processes

##### Literature gap

Although the literature provides information on the data sources that are used in priority-setting, the processes and methods used in analyzing the data are not discussed.

##### What this study adds

GHS uses the routine surveillance data in DHIMS2 for prioritization [Interviews 2, 5, 8]. PPME is responsible for managing and analyzing the data in DHIMS2 [Interviews 2, 4]. Before analyzing the data, PPME checks its accuracy, timeliness, and completeness by verifying with facilities or health directorates at multiple levels [Interview 2].

Within the health directorates, data management is led by health information officers who represent the PPME at their levels [Interview 2]. After validation, the data is analyzed in some form, using methods such as trend analysis, spot mapping using GIS, graphs, or pie charts [Interview 2]. Examples of aggregated data which inform priorities include disease burden data, health service utilization data, and demographics [Interviews 2, 3].

The health indicators considered in an analysis are dependent on the objectives of an assessment or the requester; for example, prior to certain meetings, the GHS Director may request analyses on health indicators which are relevant to the meeting theme [Interview 2]. Good coordination between the PPME and the MoH enables the MoH to present routine, up-to-date data in specific meetings while also allowing thelJPPME to stay informed of the MoH’s priorities and targets [Interview 2].

The RDD of the GHS also conducts research to inform GHS priorities [Interview 3]. Local academic institutions including Kwame Nkrumah University of Science and Technology, University of Ghana School of Public Health and University of Health and Allied Sciences, in partnership with the GHS, also conduct research to support priority-setting [Interviews 2, 6]. Research outputs include publications, policies for health promotions, policy briefs, and dialogues [Interview 2].

Given that DHIMS2 is not publicly accessible, researchers external to the GHS and commercial agencies need to request access to the database [Interviews 2, 3].

### Data and Analytic Capacity

#### Literature gap

No literature source examines the local data and analytic capacity for HSPS in Ghana. Specifically, information is absent on the gaps in local capacity, on efforts aimed at addressing these gaps and the areas these efforts are focused.

#### What this study adds

Despite the richness of data that the Government of Ghana has, its capacity to utilize and analyze the data and transform analysis results into action through policy is still very limited [Interview 7]. A lack of demand for advanced analytics (e.g., assessing relationships as opposed to simply observing counts and averages), a shortage of highly skilled professionals in data analytics, and high staff turnover exacerbates this gap [Interview 3, 4].

To address these challenges, several international agencies have been assisting Ghana in improving skills across the data life cycle. In data collection, PATH experts help to identify gaps and support the MoH to strengthen the government’s data system by improving data capture tools; for example, supporting with the addition of health indicators to the system [Interview 1]. Additionally, the WHO, as one of Ghana’s primary international partners, plays a key role in data generation for priority-setting through improvement of the SCORE assessment [Interview 7]. It also assists the government in harmonizing and conducting health facility assessments at the district level, to aid priority-setting within districts [Interview 7]. The WHO also supports in the conduct of the STEPS survey (STEPwise approach to non-communicable disease risk factor Surveillance) and the DHS which helps to generate relevant demographic data useful for health planning [Interview 7]. WHO support extends to specialized areas, such as capacity building within the tuberculosis program to improve modeling for TB incidence estimates, thereby improving the country’s disease burden assessments [Interview 7]. The WHO also conducts training for government officials and MoH stakeholders on data analysis, emphasizing the use of data for policy briefs and infographics [Interview 7]. This is intended to bolster their ability to interpret analysis results, identify programmatic gaps, and promote data-driven decision-making. Young faculty members also receive training from the WHO in implementation research to align their work more closely with national policy needs, strengthening the role of academic institutions in evidence-based policymaking [Interview 6].

Other than the WHO, other international agencies also support capacity-building efforts, with the flexibility to determine the specific level of capacity they aim to strengthen. However, at the national level, the PPME division determines the training content [Interview 2]. Ideally, such training sessions occur twice a year, though the actual frequency may vary based on funding and feedback from routine supervision [Interview 2]. Various capacity-building trainings have been conducted in 2024, such as trainings on DHIMS2 data quality, data management, standard operation procedure, district health system functionality assessment, harmonized health facility assessment, and health equity assessment [Interviews 2, 7]. By overseeing training content, PPME ensures alignment with the skills most needed by staff. For instance, DHS training specifically trains on using access mode and Quantum Geographic Information System (QGIS) for improving accessibility of emergency referrals from community-based health planning services [Interview 7].

Through capacity-building, the Information and Communication Technology department of the PPME ensures that officers at various levels can utilize data collection platforms optimally [Interview 2]. The department provides e-learning programs that have reached over 14,000 health sector staff within six years of implementation, covering various skills such as training in digital tools and descriptive analysis [Interview 2]. Training is also conducted when new features are added to DHIMS2, to ensure the quality of data entered [Interview 2]. There are efforts to train staff in predictive and prescriptive analysis [Interview 2].

### Theme 4: Successes and Challenges

There are several good aspects of HSPS in Ghana as well as gaps which need to be addressed. These are discussed in detail in this section and summarized in Fig. 2.

**Fig. 2.** Summary of successes and challenges in the health sector priority-setting process in Ghana. HSPS = Health sector priority-setting. GHS = Ghana Health Service.

### Successes

#### Priority-Setting Processes

The health sector reforms in Ghana have significantly strengthened the HSPS process, making it both more institutionalized and responsive to the country’s diverse health needs. A key reform in Ghana was decentralization, which empowered communities to participate in setting health priorities. The priorities established at the sub-national level also serve to guide those set by the GHS and the MoH at the national level, bridging the gap between the local needs and national strategies, fostering a system that is more connected and less isolated from the realities faced by communities [Interviews 1, 5, 7]. The bottom-up approach is important for Ghana’s HSPS process [Interview 1], where sub-national insights inform national strategies. The Ghanaian government has demonstrated political commitment to priority-setting.

The government has established a structured planning and budgeting cycle, with key stakeholders meeting annually during the health summit to set goals, review progress, and adjust as necessary [Interviews 1, 2, 5, 7]. This structure brings together government officials, donors, and other partners, fostering a collaborative environment that is essential for aligning priorities and ensuring accountability.

#### Data and Analytics for HSPS

DHIMS2, the main data repository for GHS facilities, consolidates information from routine health activities, creating a reliable resource for decision-making (Interview 2). In 2016/2017, Ghana introduced tablets for data collection at the community level (Interview 5), enabling seamless transfer of data to the national level thus ensuring that local insights shape national health policies in a timely fashion.

The country’s capacity to set data-driven health priorities has evolved at both the national and program-specific levels, enabling health facilities to contribute essential evidence for the development of meaningful policies. For example, within the TB program, tools developed for TB screening has improved diagnostic accuracy, while new models being developed to estimate TB incidence will provide a clearer picture of the disease burden across Ghana [Interview 8]. These advances mean that health priorities are based on current information on the health needs of the country.

Ghana has also invested in its health workforce by implementing an e-training program. Over the past six years, more than 14,000 health workers have participated in the program, learning how to collect and analyze data with precision [Interview 2].

### Challenges

#### Priority-Setting Processes

Despite Ghana’s advances in HSPS, the health sector faces significant challenges in setting these priorities effectively. Political interference often complicates the HSPS process, as decisions are sometimes swayed by shifting political agendas rather than the health needs of the population [Interviews 1, 3, 5]. This can result in disruptions in the continuity of health initiatives, especially when new political administrations, typically constituted every four years after an election, form new priorities. The influence of political cycles and competing interests make it difficult to maintain a consistent, evidence-based approach, ultimately affecting the sustainability of health policies and programs. Interviewees also cited apathy by leadership at the national level as an impediment to effective HSPS [Interviews 5, 7].

Donor influence plays a considerable role in shaping health priorities. While donor funding has been instrumental in supporting various health initiatives, it often comes with specific requirements or preferences. Donors may prioritize certain diseases or interventions that meet their organizational objectives or reflect international health trends [Interviews 2, 3, 5, 8], which could divert focus and resources from other urgent health issues in Ghana. However, development partner representatives from WHO and PATH stated that their activities align with national priorities [Interviews 1, 7]. The WHO representative also stated that its priorities are based on country data [Interview 7]. They explained that there are usually no tensions between WHO and the government with respect to what the priorities should be; however, the order of priorities usually requires discussions before agreement is reached between the parties [Interview 7]. Nevertheless, the reliance on external funds creates a complex dynamic where the government must balance donor expectations with national priorities, which may lead to uneven distributions of resources and a lack of focus on locally relevant health challenges.

Insufficient funding to implement identified priorities could be a demotivating factor for engaging in priority-setting [Interview 7]. Funding for implementation is typically limited or allocated only to specific areas; therefore, priorities without dedicated financial backing are often delayed or abandoned [Interviews 3, 5, 7]. Requested funds for project implementation are seldom available to the GHS because the Ministry of Finance, in charge of disbursing health sector funds, usually prioritizes staff salaries [Interview 5]. These make up about 70% of the funds allocated to the health sector, leaving only about 30%—an insufficient fraction—for project implementation across over 23 MoH agencies [Interview 2]. The TB Control Program within the GHS, for example, is underfunded by 45% [Interview 8]. This means that some health programs, although identified as a priority, may never be implemented if they do not attract donor interest or government allocation, limiting the impact of Ghana’s strong evidence-based planning frameworks. Indeed, an interviewee noted that research findings are often left on the shelves and inform policies when it has a lot of funding to back it [Interview 3].

Adding to the funding complexities, the separation between the MoH’s Research Statistics Information Management and the GHS’ RDD limits collaboration. Research is a significant avenue for funding, leading the MoH departments to pursue an independent agenda rather than working in collaboration [Interview 3]. High staff turnover among government officials further complicates the situation, as frequent changes disrupt continuity and institutional memory within the health sector [Interview 3].

#### Data and Analytics for HSPS

Although the Health Facilities Regulatory Agency mandates all health facilities to report health service data to the DHIMS2, teaching hospitals and some private facilities do not report their data [Interviews 2, 3, 4]. Given that these systems are not integrated [9], this situation creates a fragmented health data management system, limiting the opportunities to effectively use this data for setting priorities that addresses health issues nationally. As of August 2024, about 400 health facilities were not reporting to the DHIMS2 [Interview 4].

Limited funding hinders the operability of health databases in several aspects. Without funding for good internet connectivity, the ability to use the DHIMS or e-Tracker is limited as these databases are online [Interview 2]. Financial constraints also have prevented the full migration from the paper system to the electronic system for data management, leading to an increased workload for staff who need to operate both systems [Interview 2]. For example, of the 2000 Directly Observed Therapy centers for TB treatment, only 425 facilities—less than a quarter—report their data electronically as of August 2024 [Interview 8]. Insufficient funding also limits the feasibility of conducting routine research on the HIS, which is vital for its quality improvement [Interview 2]. Research with the HIS data is also insufficiently funded. Therefore, evidence suited to the local context is not always available, causing stakeholders to adopt policies based on evidence from other countries [Interview 3, 4]. Such policies are likely to fail, given that context-specific information impacts generated evidence.

Additionally, the lack of demand for more advanced analytics, such as predictive analysis, prescriptive analysis, big data analysis, data visualization, analysis of relationships between factors, creates a disincentive for current GHS staff to pursue further training in advanced data analysis and for doctoral level data analysts to work with the GHS (Interview 3). The main type of analysis used in MoH and GHS reports are descriptive analysis such as counts, averages and pie charts [Interviews 2, 4]. This limitation increases the government’s reliance on external support for analytical needs [Interview 4].

### Recommendations for Improvement of HSPS

The following recommendations outline short-term and long-term actions for improvement of HSPS in Ghana. These recommendations are formulated based on information from the interviews, literature review and evidence from other countries. These recommendations are summarized in Fig. 3 and detailed below.

**Fig. 3.** Recommendations for health sector priority-setting in Ghana. HSPS = Health sector priority-setting.

### Data and Analytics

Currently, health data is not centralized within a single agency, making it imperative to enhance interoperability and integration within existing health information systems [9,21]. By connecting and coordinating these systems, health data can be used more effectively, supporting a unified approach to HSPS [Interview 4]. To achieve this, the government should assess the technical readiness of each system for integration, data accessibility, and data management. For example, assessing the effectiveness of health indicators in existing systems such as DHIMS2 [Interview 4] will be important in enabling a more accurate understanding of the national status of healthcare delivery. Strengthening data reporting regulations, through HEFRA, to promote compliance is a needed action if systems integration is to be successful in practice.

The ownership of databases is an important issue. When the software used to develop national health databases are proprietary and owned by entities other than the government, it makes the country vulnerable to decisions of owners to withhold data in the event of contractual misunderstandings. For example, an alleged delay in government’s payment for the services of LHIMS, which is privately owned, has raised concerns about possible leaks of sensitive private health data as a bargaining tool [22]. Additionally, the recent cuts in USAID funding have led to restricted access to DHS data, limiting the opportunities for research and interventions on population health [23].To preserve patients’ privacy and to ensure the availability of critical public health data, any national database should be owned and managed by the government.

Additionally, the government should establish clear health data management policies and strategies that define the roles and responsibilities of every related actor and how actors can coordinate with one another in data sharing and data access [24–26]. Involvement of leadership from teaching hospital and private sector facilities in HIS planning and coordination could engender a greater sense of ownership of data in the health sector, to address the perception of teaching and private facilities that they are not part of the district health system [9].

The MoH should establish one national health research agency––such as exist in Tanzania with the National Institute for Medical Research, in Kenya with the Kenya Medical Research Institute, and in South Africa with the South African Medical Research Council––to reduce fragmentation and duplication of efforts between the MoH’s Research, Statistics and Information Management Directorate and the GHS’ RDD, and to strengthen the coordination, performance, and capacity of health research in Ghana. Given that the MoH is the policy-making body, it would be beneficial to position this agency within the MoH [Interview 3].

Integrating the country’s expertise, resources, and data into one health research agency would address challenges related to human resources, institutional capacity, absence of clear coordination mechanisms, and fragmentation in funding for health research. This would strengthen the use of data from in-country research to inform HSPS.

Building a culture of training in, and demand for, data-driven decision-making is crucial to improving data use and analysis within the health sector. One method is to first conduct an analytic capacity needs assessment [Interview 4]. Based on the assessment, training programs aimed at developing advanced analytical skills could be designed. In addition to presenting or visualizing data effectively [Interviews 2, 4], these programs can focus on developing complex critical thinking skills [Interview 3], such as identifying relationships between factors [Interview 4], enabling staff to perform more in-depth analyses to generate insights which would have not been possible with basic descriptive analyses [21]. While Ghana has established a program to train personnel at lower levels of the health system in data analytics, expanding training to senior staff and directors, including within the MoH, would be beneficial [Interviews 3, 4] as this group more directly impact the final stages of priority-setting. Capacity improvement is needed not only in data analysis, but also in data generation. For example, to improve TB case detection, capacity improvement is needed among laboratory scientists, nurses and referral clinicians [Interview 8]. Another avenue to increase data proficiency across the health workforce could be task shifting; for example, a nurse learning how to input data so they can do so in the event of unavailability of a data manager [Interview 8]. Achieving this effectively would require an expansion in the training curricula of nurses and other allied health professionals, to include data analytic content. Importantly, one interviewee also emphasized that expanding capacity building training programs alone is insufficient for staff to advance their analytic skills; programs must be complemented with a greater culture of demand for the use of advanced analytics to motivate staff, as well as to improve the attraction and retention of qualified statisticians from the academic or private sectors to the MoH [Interview 3].

The Individuals, Nodes, Networks, and Environment framework could serve as a valuable model for planning these capacity-building efforts. The framework emphasizes locally relevant technical skills in each stakeholder’s specific environment, maximizing the application of skills throughout the priority-setting processes [27]. In addition, the government could introduce performance indicators related to evidence use, such as the integration of research into policymaking and the application of skills gained through training in data analytics.

Incorporating these indicators within performance assessments for directors in the national level agencies could facilitate the adoption and effectiveness of capacity-building programs in data and analytics [21].

### Funding Sustainability in Information Systems

In Ghana, the main health information system—DHIMS2—is heavily supported by external donors [Interview 2; 28]. For Ghana’s health sector to secure long-term sustainability, the government must provide comprehensive monitoring and evaluation reports, demonstrating the impact and effectiveness of the system [29]. Evidence can help convince current donors to maintain their support and potentially attract new investors. Given that it is possible for donors to withdraw their entire support, as seen with USAID, the past principal funder of e-Tracker [28], better sustainability would be achieved by committing local funds to support these systems.

This commitment involves a well-defined national HIS plan that includes a solid funding strategy, potentially leveraging public-private partnerships, donor contributions or allocations from the national budget [30]. Moreover, to strengthen the financial support for the sustainability of the information system, the government can collaborate with private sector financing and build strong multisectoral partnerships to increase resources.

### Priority-Setting Processes

While the government of Ghana has utilized data to inform its HSPS process, it lacks clearly defined criteria such as cost-effectiveness, health benefit, or equity, that can provide structure and transparency [31]. Establishing clear criteria would help guide stakeholders, ensuring that the prioritization process aligns with the local values and context of the country [32]. Additionally, subnational governments and communities should also be informed of these criteria, so they can better participate in the process, even appealing national decisions that are inconsistent with the criteria.[33] Such improved participation could lead to a sense of ownership of decisions among subnational health directorates, thereby decreasing the observed “disconnect between local and national levels,” where national priorities are not implemented properly at subnational levels [Interview 5].

Limiting donor influence on HSPS will also require that funding for healthcare is increasingly sourced locally. The success of a long-term domestic financing plan for healthcare in Ghana will be contingent on a strong national commitment, to ensure that the plan is still implemented even if political leadership is changed. Improvement of local funding will also aid in addressing the demotivation associated with setting priorities that will not be implemented due to lack of funds.

A closer collaboration between wider sectoral and program-specific priority setting will improve the harmonization towards the achievement of health sector goals and may improve efficiency in resource use. However, such collaboration may be challenging for donor-funded programs, if the donor does not deem collaboration a priority [Interview 8].

As only a few government representatives, including the Minister of Health and some directors at the GHS and MoH, participate in final priority-setting at the national level [Interviews 1, 5], it is critical to strengthen the decision-making capacity of these individuals. To do this, the government should establish a core group of these key representatives and design capacity-building initiatives for this group [Interview 4], thus enabling them to make well-informed decisions, improving the overall effectiveness of HSPS.

## Discussion

Using interviews with key informants and qualitative analysis, this study addressed important gaps identified in a literature review on HSPS in Ghana. Specifically, the paper provided an in-depth examination of the institutions involved in Ghana’s HSPS; the health assessments which inform the final set of priorities; the processes between the conduct of assessments and formation of priorities; data and analytics used in HSPS and the successes and challenges associated with HSPS. Based on the detailed literature review, informative interviews and evidence from other countries, the paper also offers recommendations for the improvement of HSPS in Ghana.

The idea that was reinforced in many of the interviews was that prioritization in Ghana’s health sector is heavily influenced by the availability of resources rather than by objective criteria. Due to a high reliance on development partner support for implementation of priorities, HSPS in Ghana is heavily influenced by donor priorities [Interviews 2, 3, 5], creating a situation where priorities may not reflect the country’s health needs. There was a disagreement between interviewees on this point, with two development partner representatives—from WHO and PATH—claiming that their organizations’ activities align with government’s priorities [Interviews 1, 7]. Political influences were also cited as an impediment to effective HSPS [Interviews 1, 5]. These influences could be ameliorated by establishing clear objective criteria for HSPS to help guide stakeholders and by empowering the public to contribute to setting the criteria while holding the government accountable to the set criteria. Committing to the generation of local funds for financing the implementation of priorities is a needed step if donor influences on health sector decisions are to be minimized.

The analysis revealed that though the MoH leads in priority-setting, it relies mainly on assessments conducted by the GHS, its implementation agency. GHS assessments are mainly based on the DHIMS2 which is separate from health databases used by teaching hospitals and some private health facilities [Interviews 2, 3]. Both the literature review and the interviews showed that the country does not have a unified health service database, and that there is limited interoperability between the various health data systems [Interviews 2, 3; 9]. Sustainability of HIS in Ghana could be achieved by creating long-term strategies for HIS and collaborating with in-country private sector for financing, rather than relying on external donor funds which could be unstable and associated with conditions.

The analysis method used for an assessment depends on the purpose of the assessments but are mostly counts, averages and charts [Interviews 2, 4], though there is a desire to use more advanced analytics [Interviews 2, 3, 4] and data visualization [Interview 2]. Multiple interviewees emphasized that the issue is not a lack of data; rather, the main challenge lies in capacity to synthesize, analyze, and translate the data into actionable policy [Interviews 2, 4, 7]. Indeed, the interviews revealed efforts being made to develop data and analytic capacity within the health sector [Interviews 2, 6, 7, 8], such as training of MoH representatives on the use of data for policy briefs and infographics [Interview 7]. However, many of these efforts are targeted at individuals who are not involved in priority-setting at the highest level, limiting their influence on the latter, yet critical stages of HSPS.

While this study sought to address the gaps identified in the literature review on HSPS, this study was limited by its sampling strategy. Purposive sampling makes it challenging to evaluate the generalizability of the findings here, given that interviewees were selected in a non-random fashion. However, achieving saturation provided assurance that key information regarding HSPS in Ghana had been sufficiently covered by the interviews. Also, given that interview responses were recorded by notetaking and not verbatim, it is possible that interviewers may have missed important details. To minimize errors due to notetaking, notes were taken by two interviewers and were compared to achieve consistency. Again, despite efforts to ensure objectivity, researcher biases may have subtly influenced the interpretation and presentation of findings. Efforts have however been made to present findings transparently, with a ten-member research team informing analysis, interpretation and presentation of results over several iterations.

Despite these weaknesses, the study has several strengths, key of which is the use of both an in-depth literature review and interviews with key informants to present a broad and current view of HSPS in Ghana with a focus on data and analytics. Another strength is the background and extensive experience of interviewees, which allowed the leveraging of institutional memory for the realization of useful insights, many of which were absent from the literature. The study fills important gaps in the understanding of HSPS in Ghana and provides a good foundation for further study on the subject.

There are many potential areas for further study. This work has focused on HSPS at the national level. Further study could explore HSPS at the subnational level, and the opportunities for improvement of engagement between stakeholders at the national and sub-national levels. Another potential area for further study could be an analysis of existing efforts on the recommendations offered in this paper; for example, the recommendation to integrate health data management systems, and an associated examination of factors required to effectively implement those recommendations for improved HSPS in Ghana. Multiple interviewees cited implementation of priorities as a major issue for the health sector in Ghana [Interviews 4, 5, 6]. It is therefore worth studying the barriers to program implementation, especially the role of health sector financing.

## Conclusion

This study sought to understand the use of data for HSPS in Ghana using qualitative key informant interviews to fill gaps in the literature. The Ghanaian government has demonstrated political commitment to priority-setting with an established planning and budgeting cycle and attempts at a bottom-up approach to priority-setting. However, interviewees highlighted challenges with Ghana’s HSPS processes, emphasizing that priority-setting is influenced by donor interests, political interference, a shortage of local funding or resources to implement priorities, weaknesses in HIS infrastructure and an insufficient capacity and lack of demand for advanced data analytics. Recommendations for improvements of Ghana’s HSPS processes include developing a nationally owned, sustainable and unified HIS, developing capacity for data and analytics for decision making and committing to a long-term domestic financing plan for healthcare.

## Data Availability

All associated data have been included in the supporting files.

## Acknowledgements

We thank interviewees for sharing their experiences based on their work at the Ministry of Health, Ghana Health Service, WHO Ghana, PATH Ghana and the University of Health and Allied Sciences. We also thank Dr. Isaac Agorinya and faculty at the Fred N. Binka School of Public Health, University of Health and Allied Sciences, for their input.

## Disclaimer

Interviewees shared their own insights; they did not speak for their organizations.

## Supporting information

**S1 Text. Further information from the literature on HSPS in Ghana.**

**S2 Text. Interview guides.**

**S3 Text. Interview notes.**

## References

1. Republic of Ghana. Ghana Health Service and Teaching Hospital Act [Internet]. Act 525 1996. Available from: https://lawsghana.com/post-1992-legislation/table-of-content/Acts%20of%20Parliament/GHANA%20HEALTH%20SERVICE%20AND%20TEACHING%20HOSPITALS%20ACT,%201996%20(ACT%20525)/138

2. Ministry of Health Republic of Ghana. National Health Policy: Ensuring health lives for all (Revised Edition) [Internet]. Ministry of Health Republic of Ghana; 2020. Available from: https://www.moh.gov.gh/wp-content/uploads/2021/08/NHP_January-2020.pdf

3. UNICEF. Ghana’s Health Budget Brief 2023 [Internet]. UNICEF; 2023. Available from: https://www.unicef.org/ghana/media/5001/file/2023%20Health%20Budget%20Brief.pdf

4. Ministry of Health Republic of Ghana. Medium Term Expenditure Framework (MTEF) for 2024 - 2027 [Internet]. Ministry of Health Republic of Ghana; 2024 [cited 2025 Dec 1]. Available from: https://mofep.gov.gh/sites/default/files/pbb-estimates/2024/2024-PBB-MoH_.pdf

5. Kates J, Rouw A, Oum S. U.S. Foreign Aid Freeze & Dissolution of USAID: Timeline of Events [Internet]. KFF. 2025 [cited 2025 May 29]. Available from: https://www.kff.org/u-s-foreign-aid-freeze-dissolution-of-usaid-timeline-of-events/

6. Schmets G, Rajan D, Kadandale S, editors. Strategizing national health in the 21st century: a handbook [Internet]. Gevena: World Health Organization; 2016 [cited 2025 May 19]. 712 p. Available from: https://www.who.int/publications/i/item/9789241549745

7. Baker P, Barasa E, Chalkidou K, Chola L, Culyer A, Dabak S, et al. International Partnerships to Develop Evidence-informed Priority Setting Institutions: Ten Years of Experience from the International Decision Support Initiative (iDSI). Health Syst Reform. 2023 Dec 31;9(3):2330112.

8. Koduah A, Anim Boadi J, Azeez JNK, Adu Asare B, Yevutsey S, Gyansa-Lutterodt M, et al. Institutionalizing Health Technology Assessment in Ghana: Enablers, Constraints, and Lessons. Health Syst Reform. 2023 Dec 31;9(3):2314519.

9. Ministry of Health Republic of Ghana. Health Information System Strategic Plan 2022-2025. 2022.

10. Koduah A, Anim Boadi J, Azeez JNK, Adu Asare B, Yevutsey S, Gyansa-Lutterodt M, et al. Institutionalizing Health Technology Assessment in Ghana: Enablers, Constraints, and Lessons. Health Syst Reform. 2023 Dec 31;9(3):2314519.

11. Birungi H, Nyarko P, Askew I, Ajayi A, Addico G, Addai E, et al. Priority setting for reproductive health at the district level in the context of health sector reforms in Ghana. Reprod Health [Internet]. 2006 Jan 1; Available from: https://knowledgecommons.popcouncil.org/departments_sbsr-rh/398

12. Saunders B, Sim J, Kingstone T, Baker S, Waterfield J, Bartlam B, et al. Saturation in qualitative research: exploring its conceptualization and operationalization. Qual Quant. 2018;52(4):1893–907.

13. Ministry of Health Republic of Ghana. Health Sector Medium Term Development Plan 2022-2025 [Internet]. Ministry of Health Republic of Ghana; 2021 [cited 2024 Sep 16]. Available from: https://www.globalfinancingfacility.org/resource/ghana-health-sector-medium-term-development-plan-2022-2025

14. Ghana Health Service. GHS Governance System [Internet]. Ghana Health Service. [cited 2024 Oct 1]. Available from: https://ghs.gov.gh/ghs-governance-system/#:~:text=Ghana%20Health%20Service%20Council,controlling%20the%20affairs%20of%20GHS

15. Hollingworth SA, Downey L, Ruiz FJ, Odame E, Dsane-Selby L, Gyansa-Lutterodt M, et al. What do we need to know? Data sources to support evidence-based decisions using health technology assessment in Ghana. Health Res Policy Syst. 2020 Apr 28;18(1):41.

16. Ministry of Health Republic of Ghana. Health Sector Annual Programme of Work - 2022 Holistic Assessment Report [Internet]. Ministry of Health Republic of Ghana; 2021 Apr p. 65. Available from: https://www.moh.gov.gh/wp-content/uploads/2022/09/2020-Holistic-Assessment-Report_v8.3docx.pdf

17. Ministry of Health Republic of Ghana. Ghana Health Sector 2024 Programme of Work [Internet]. Ministry of Health Republic of Ghana; 2024. Available from: https://www.moh.gov.gh/wp-content/uploads/2024/04/Final-2024-Annual-Programme-of-Work.pdf

18. Ministry of Health Republic of Ghana. Common Management Arrangements for Implementation of the Health Sector Medium-Term Development Plan (2022-2025) [Internet]. Ministry of Health Republic of Ghana; 2023. Available from: https://www.moh.gov.gh/wp-content/uploads/2025/01/CMA-2022-2025_FINAL.pdf

19. Ministry of Health Republic of Ghana. Health Sector Annual Summit 2024 [Internet]. Ministry of Health Republic of Ghana. 2024 [cited 2024 Oct 30]. Available from: https://www.moh.gov.gh/heath-summit-2024/

20. Ministry of Health Republic of Ghana. Ghana’s Health Sector Annual Summit 2024: Quality Healthcare Delivery, A Catalyst for Achieving Universal Health Coverage in Ghana [Internet]. Ministry of Health Republic of Ghana; 2024. Available from: www.moh.gov.gh/wp-content/uploads/2024/05/HEALTH-Summit-Program-2024-NEW.pdf

21. Joint Learning Network for Universal Health Coverage. Measuring The Performance of Primary Health Care: A Practical Guide for Translating Data into Improvement. Joint Learning Network for Universal Health Coverage (JLN); 2018.

22. Data at Risk: Inside the Ministry of Health’s controversial $100m partnership - Report [Internet]. GhanaWeb. 2025 [cited 2025 Jun 24]. Available from: https://www.ghanaweb.com/GhanaHomePage/NewsArchive/Data-at-Risk-Inside-the-Ministry-of-Health-s-controversial-100m-partnership-Report-1988160

23. Khaki JJ, Molenaar J, Karki S, Olal E, Straneo M, Mosuse MA, et al. When health data go dark: the importance of the DHS Program and imagining its future. BMC Med. 2025 Apr 24;23(1):241.

24. Country Health Information Systems and Data Use. Global Health Security (GHS) Surveillance Analysis and Data Use: Synthesis Report. Country Health Information Systems and Data Use; 2024 Jan.

25. Jayatissa P, Hewapathirana R. Enhancing Interoperability among Health Information Systems in Low and Middle-Income Countries: A Review of Challenges and Strategies. Eur Mod Stud J. 2023 Aug 10;7(3):334–40.

26. Koumamba AP, Bisvigou UJ, Ngoungou EB, Diallo G. Health information systems in developing countries: case of African countries. BMC Med Inform Decis Mak. 2021 Aug 4;21(1):232.

27. Li R, Ruiz F, Culyer AJ, Chalkidou K, Hofman KJ. Evidence-informed capacity building for setting health priorities in low- and middle-income countries: A framework and recommendations for further research [Internet]. F1000Research; 2017 [cited 2025 May 19]. Available from: https://f1000research.com/articles/6-231

28. Abotsi E, Afenyadu G, Yentumi G, Nyuzaghl JA, Biritwum-Nyarko A, Ofosu AA, et al. Transitioning to digital transactional data capture in primary health care facilities: a case report from Ghana’s Savannah Region. mHealth [Internet]. 2025 Jan 30 [cited 2025 May 29];11(0). Available from: https://mhealth.amegroups.org/article/view/133615

29. Akhlaq A, McKinstry B, Muhammad KB, Sheikh A. Barriers and facilitators to health information exchange in low- and middle-income country settings: a systematic review. Health Policy Plan. 2016 Nov 1;31(9):1310–25.

30. Labrique AB, Wadhwani C, Williams KA, Lamptey P, Hesp C, Luk R, et al. Best practices in scaling digital health in low and middle income countries. Glob Health. 2018 Nov 3;14(1):103.

31. Kaur G, Prinja S, Lakshmi PVM, Downey L, Sharma D, Teerawattananon Y. Criteria Used for Priority-Setting for Public Health Resource Allocation in Low- and Middle-Income Countries: A Systematic Review. Int J Technol Assess Health Care. 2019 Jan;35(6):474– 83.

32. Kapiriri L, Martin DK. A Strategy to Improve Priority Setting in Developing Countries. Health Care Anal. 2007 Sep 1;15(3):159–67.

33. Kwete XJ, Berhane Y, Mwanyika-Sando M, Oduola A, Liu Y, Workneh F, et al. Health priority-setting for official development assistance in low-income and middle-income countries: a Best Fit Framework Synthesis study with primary data from Ethiopia, Nigeria and Tanzania. BMC Public Health. 2021 Nov 21;21(1):2138.

