## Supplementary material for "The use of data and analytics for health sector priority-setting in Ghana: a qualitative study": S1 Text

**S1 Text: Further information from literature review**

### Health Governance Structure

Figure A. GHS governance system.

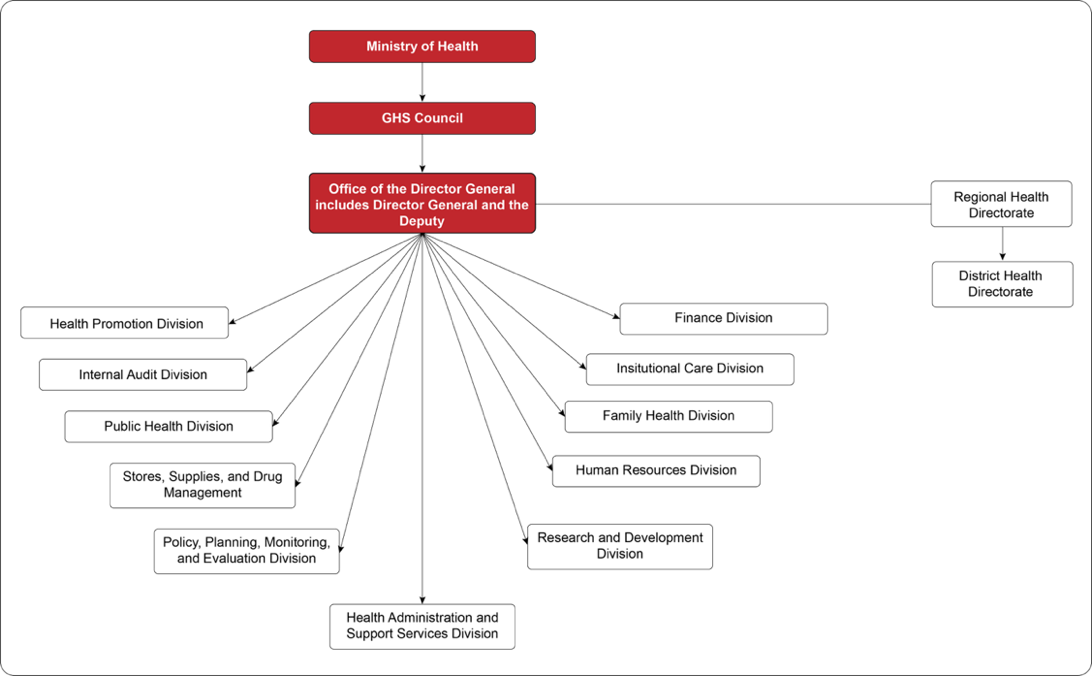

Note: GHS = Ghana Health Service. Source: Adapted from [1].

Figure B. Structure of Ghana's health system.

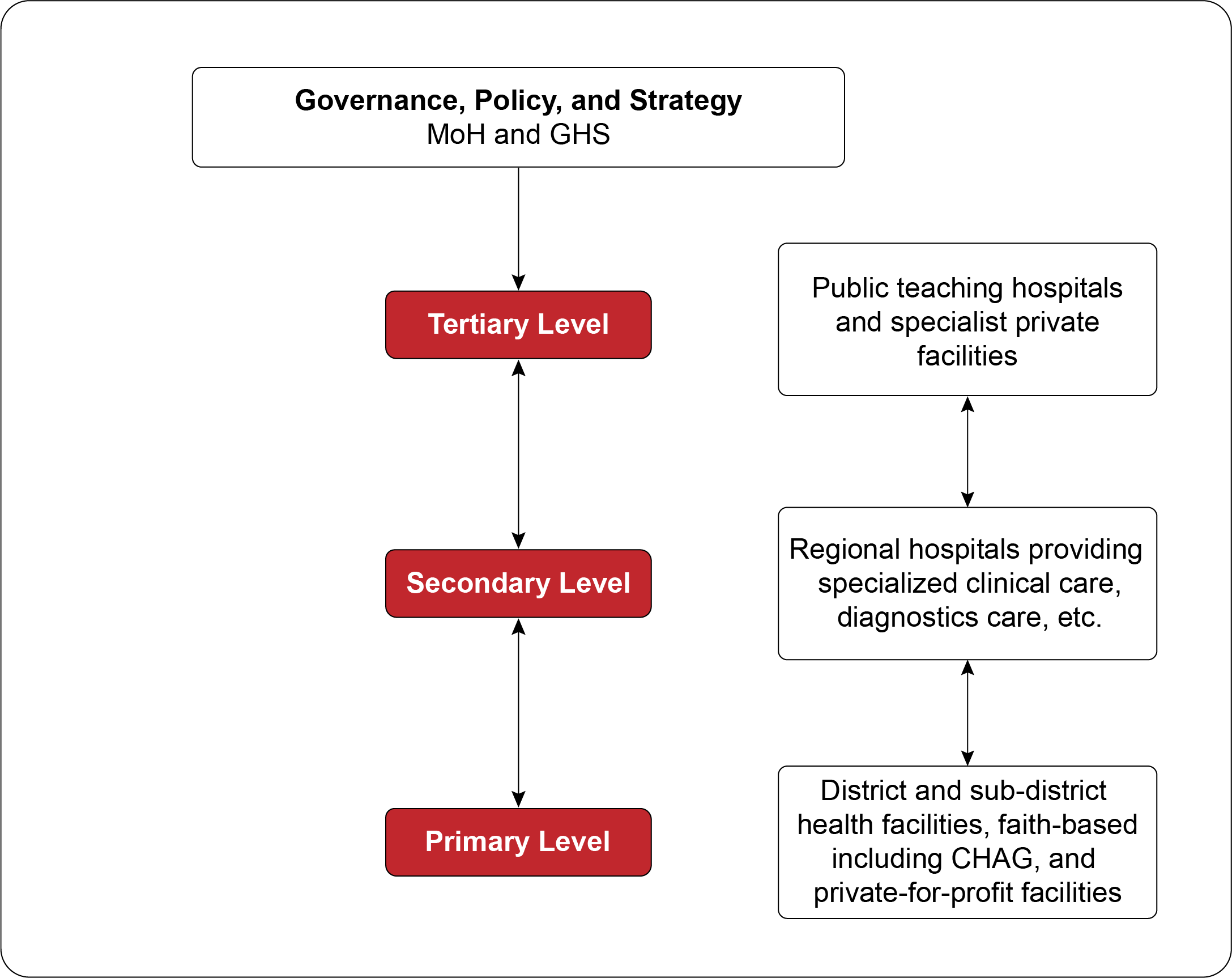

Note: MoH = Ministry of Health, GHS = Ghana Health Service, CHAG = Christian Health Association of Ghana. Source: Adapted from [2].

### Health Service Data

#### Data Sources

Ghana has several health data sources, such as the Demographic and Health Survey (DHS) data, Global Burden of Disease (GBD), National Health Insurance Scheme (NHIS) data, Human Resource Information System (HRIS) for the health sector, and the District Health Information Management System (DHIMS2) which is the main repository of health service data for the GHS and MoH [3,4].

The DHS data is based on a nationally representative household population survey conducted every five years on demographic and health indicators including maternal and child health, fertility, sanitation, and standard of living. Detailed raw data from this survey can be accessed by request, but the report is freely available online [5]. Data from NHIS claims can be used to assess health service use, medicine costs, and tracking performance of the schemes, but it has limited access [3]. Using national datasets from DHIMS and stakeholder consultation, Ghana validates the GBD country-specific information. This updated information is later used to update the National Health Policy, the HSMTDP, and the UHC roadmap [3].

Figure C shows the data flow in the Ghana Health Information System (HIS), capturing the input and feedback flows from various health facilities and governance levels. The figure shows that there are multiple health care data reporting platforms within the health sector, with separate electronic health record (EHR) systems for teaching hospitals, quasi-government facilities, and some private hospitals that are not integrated in the GHS’ DHIMS2. Facilities are to report health care data through the District/Sub-Municipal Health Directorates to collate and analyze within the DHIMS2. However, these platforms often have limited interoperability, leading to incompleteness in service delivery reports in districts with these facilities. Additionally, there is the absence of formal inter-agency coordinating mechanisms for health information [4].

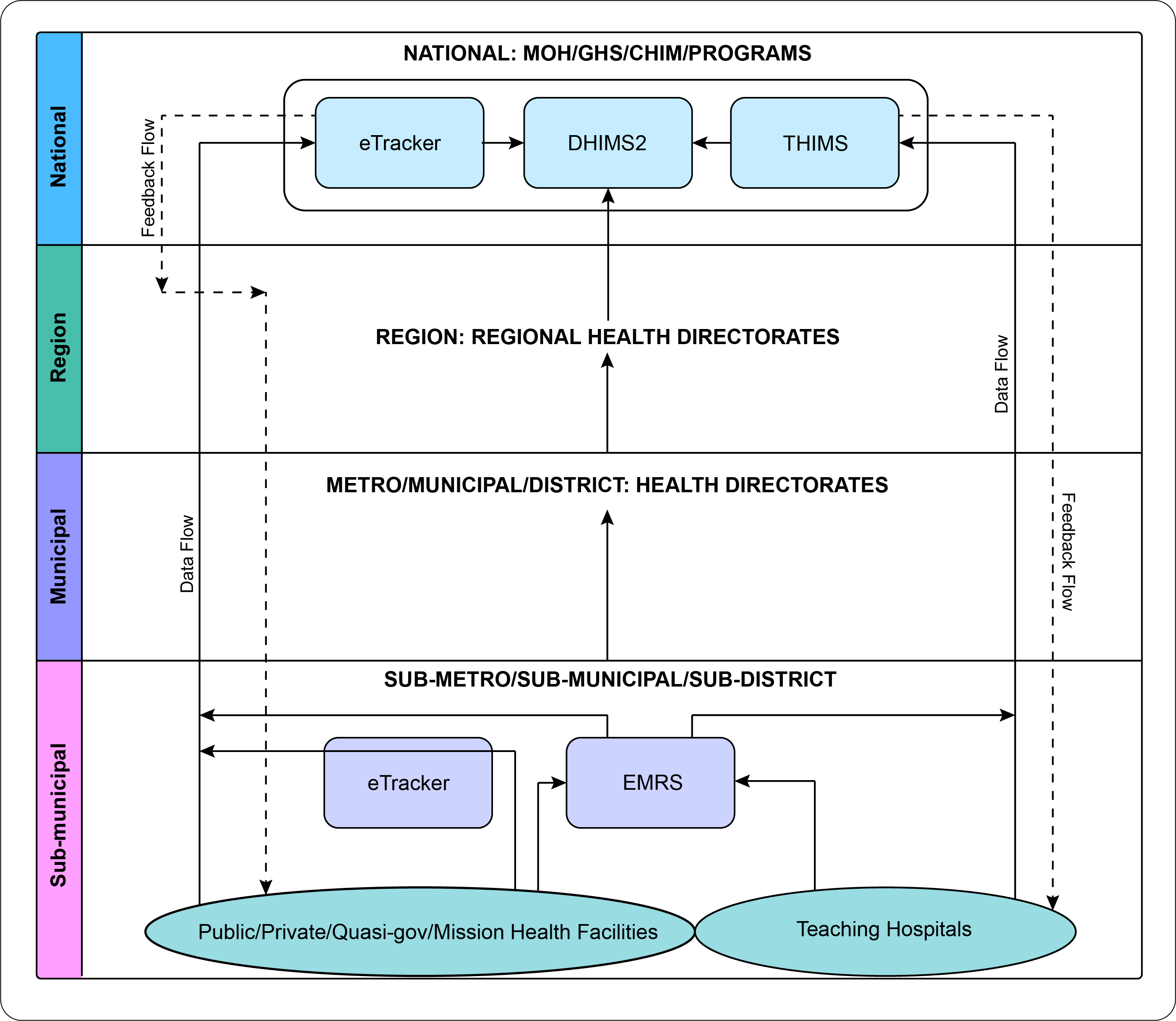

Figure C: Data flow in the health information system. Note: MOH = Ministry of Health, GHS = Ghana Health Service, CHIM = Centre for Health Information Management, DHIMS2 = District Health Information Management System 2, THIMS = Teaching Hospitals Information Management System. Source: Adapted from [4].

##### DHIMS-2

*Structure and function.* The DHIMS2 is the implementation of District Health Information System (DHIS), a web-based database that manages routine health service data from health facilities under the GHS [4]. DHIMS2 allows reporting on various variables including care delivery, public health events, catchment areas of health facilities, human resources, equipment, and facility capacity [6].

The DHIMS2 platform uses a cloud server-based architecture to standardize data systems across all health facilities, enabling analysis and comparison of datasets at individual or multiple health facilities. Additionally, DHIMS2 offers various visualization options, ranging from simple bar graphs to geo-location-based visualization, which encourages facilities to increase information uptake [6]. During the COVID-19 pandemic, data from DHIMS2 supported the disease surveillance framework to assist the decision-making process, especially in capturing vaccination coverage in the population [7].

*Access.* As the DHIMS2 can only be accessed directly by the staff of the MoH and its agencies, other organizations can request data from the Director General of the GHS, through the PPME Directorate.

*Challenges.* While DHIMS2 has proven effective in many areas, it faces significant human resources, logistics, and data quality challenges. Data accuracy remains a concern, due to limited training for data collectors, gaps in data capture, infrequent verification, and inconsistencies in the coding of health indicators [8]. Furthermore, the national-level periodic shutdown of DHIMS2—intended to verify data at the district level—may inadvertently prompt health facilities to over-report specific indicators [9]. Recognizing these challenges, the GHS has worked actively with partner agencies to establish robust data quality improvement measures. These efforts focus on standardizing procedures and outputs to achieve reliable, high-quality data [8,10].

The scarcity of logistics, such as inadequate functional computers and unstable internet access in some regions, influenced user’s acceptance of the DHIMS because it is internet-dependent [11]. In some cases, the mental health condition of healthcare workers may cause them to neglect the reporting of a particular type of data. For example, trauma and guilt associated with stillbirth can cause underreporting by healthcare workers [9]. Furthermore, the health workforce's limited skillset is further impacted by the lack of a comprehensive reference manual to guide users on the DHIMS2 platform, as well as infrequent training opportunities at the district and sub-district levels [6]. These limitations hinder the effective use of evidence-based data in decision-making processes at the community and sub-district levels [6].

##### e-Tracker

*Structure and function.* E-Tracker is an Electronic Health Records (EHR) system and a part of the individual client-based module in the DHIMS2 within the CHPS. It captures data on antenatal care, delivery, postnatal care, and family planning services [12]. Initially implemented in public health facilities at the district level, the e-Tracker was scaled up by the GHS to the community level in 2014 and eventually deployed nationwide [12]. The collated data have helped health facilities identify missing service patients, retrieve medical records, support the continuum of care, design maternal/child/family planning interventions, procure supplies needed for maternal and child services, evaluate community health nurse (CHN) performance, provide evidence for the CHPS compound, and write research proposals on maternal and child health [11–14]. The tablet-based electronic application also streamlines data validation. It generates statistics on vaccination and antenatal coverage, although it has not significantly enhanced data quality compared to the traditional paper-based system [11].

*Challenges.* Many parts of Ghana face unstable internet connectivity and inconsistent power supply, making it challenging to utilize the e-Tracker application. This negatively impacts staff acceptance of the application and often compels them to rely on their personal data packages for syncing e-Tracker information [11–14]. As a result, healthcare providers are unable to use the e-Tracker during patient care and while generating retrospective reports, which can compromise the timeliness and quality of collected data and increase the workload for staff [12]. Additionally, various challenges related to health personnel—including limited skills in tablet usage, inadequate training on the e-Tracker, high staff turnover, and perceptions of increased workload—have contributed to the lower acceptance of the application among healthcare workers, leading many to continue using paper-based reporting [12,13]. The design limitations of tablet computers can also make data entry difficult during busy periods [12]. In terms of data quality, discrepancies exist between electronic and paper-based individual records, primarily due to missing values in certain data elements that lack validation rules [12].

##### Surveillance Outbreak Response Management and Analysis System (SORMAS)

*Structure and function.* SORMAS [15] is a digitalized health system that was first developed in the context of the 2014-2016 West Africa Ebola virus outbreak but was nationally introduced in Ghana during the COVID-19 pandemic to help the government monitor and manage the spread of the virus in communities [16]. SORMAS provides open-source data on new cases, recoveries, and deaths, accessible to the public and government. This electronic system helps healthcare workers streamline their work routines by integrating laboratory and surveillance procedures, standardizing data collection and reporting templates across the country, providing real-time reports, and supporting the government to track regional case count breakdowns [16].

*Challenges.* Despite these benefits, SORMAS faces challenges, including an overly complex interface, the lack of an offline mode of the web version, and unstable interoperability with the DHIMS2 system [7,16].

##### Lightwave Health Information Management System (LHIMS)

*Structure and function.* LHIMS is a web-based software platform that aims to facilitate the integration of healthcare data systems in Ghana [17]. It enables providers rapid access to patients’ records and real-time service information such as bed availability, thereby promoting quick data-driven decision-making [7,18,19]. LHIMS was initiated in 2018 by the MoH [17] but was officially launched in October 2024 [20] after a pilot. It is not clear from the literature whether the LHIMS is intended to replace the DHIMS2 or offer an alternative to it.

As an EHR system, it has comprehensive components that help providers submit NHIA claims, retrieve patients’ records, schedule appointments, and access advance examination results [18]. Its nature as an electronic system has improved data quality, especially in terms of timeliness, security, consistency, reliability, and accuracy [19]. The LHIMS improves the efficiency of care delivery, lessening the amount of time a patient spends at a care unit [18,19]. The real-time features of this system significantly guided the government in disease surveillance during the COVID-19 pandemic, assisting them in timely case identification and analyzing further pandemic response needed [7].

*Challenges.* However, LHIMS requires reliable internet connectivity, which is challenging in remote and low-resource settings, leading to unequal data distribution and hampering the decision-making process during outbreaks [7,19]. Insufficient logistics and technological drive and an unstable power supply, aggravates this condition, hindering providers from inputting or retrieving patients’ information [19]. Additionally, some critical patient information is not included, such as radiology and laboratory results, drug allergies, and drug dosing alerts [19]. Patient sociodemographic information is also not included in the system, affecting data-driven, informed decision-making in the public health sector [7].

#### Future Plans for Health Information Systems

Ghana’s HIS has many strengths, including the existence of DHIMS2 [4], institutionalized performance review of health information systems at the district, regional and national levels, and the existence of an office—CHIM—to administrate healthcare data management. However, many challenges remain as outlined in preceding sections, key of which is the absence of an integrated, national health database. To address these challenges, the government, under the guidance of the MoH, set reforms in the Health Information System Strategic Plan (HISSP) 2022 – 2025 with the goal to institutionalize an integrated health information system that will provide a high-quality, comprehensive national database and dashboard to support evidence-based decision-making in HSPS [4].

### References

1.	Ghana Health Service. GHS Governance System [Internet]. Ghana Health Service. [cited 2024 Oct 1]. Available from: https://ghs.gov.gh/ghs-governance-system/#:~:text=Ghana%20Health%20Service%20Council,controlling%20the%20affairs%20of%20GHS

2.	Ministry of Health Republic of Ghana. Health Sector Medium Term Development Plan 2022-2025 [Internet]. Ministry of Health Republic of Ghana; 2021 [cited 2024 Sep 16]. Available from: https://www.globalfinancingfacility.org/resource/ghana-health-sector-medium-term-development-plan-2022-2025

3.	Hollingworth SA, Downey L, Ruiz FJ, Odame E, Dsane-Selby L, Gyansa-Lutterodt M, et al. What do we need to know? Data sources to support evidence-based decisions using health technology assessment in Ghana. Health Res Policy Syst. 2020 Apr 28;18(1):41.

4.	Ministry of Health Republic of Ghana. Health Information System Strategic Plan 2022-2025. 2022.

5.	Ghana Statistical Service (GSS), ICF. Ghana Demographic and Health Survey 2022 [Internet]. Accra, Ghana and Rockville, Maryland, USA; 2024 p. 653. Available from: https://dhsprogram.com/pubs/pdf/FR387/FR387.pdf

6.	Odei-Lartey EO, Prah RKD, Anane EA, Danwonno H, Gyaase S, Oppong FB, et al. Utilization of the national cluster of district health information system for health service decision-making at the district, sub-district and community levels in selected districts of the Brong Ahafo region in Ghana. BMC Health Serv Res. 2020 Jun 6;20(1):514.

7.	Owusu I, Acheampong GK, Akyereko E, Agyei NA, Ashong M, Amofa I, et al. The role of digital surveillance during outbreaks: the Ghana experience from COVID‑19 response. J Public Health Afr. 2023 Oct 30;14(10):10.

8.	Kayode GA, Amoakoh-Coleman M, Brown-Davies C, Grobbee DE, Agyepong IA, Ansah E, et al. Quantifying the Validity of Routine Neonatal Healthcare Data in the Greater Accra Region, Ghana. PLOS ONE. 2014 Aug 21;9(8):e104053.

9.	Mensah Abrampah NA, Okwaraji YB, Oteng KF, Asiedu EK, Larsen-Reindorf R, Blencowe H, et al. District health management and stillbirth recording and reporting: a qualitative study in the Ashanti Region of Ghana. BMC Pregnancy Childbirth. 2024 Jan 29;24(1):91.

10.	Owusu MF, Adu J, Dortey BA. “I tell you, getting data for this is hell”–Exploring the use of evidence for noncommunicable disease policies in Ghana. PLOS Glob Public Health. 2023 Aug 24;3(8):e0002308.

11.	Lee S, Lee YJ, Kim S, Choi W, Jeong Y, Rhim NJ, et al. Perceptions on Data Quality, Use, and Management Following the Adoption of Tablet-Based Electronic Health Records: Results from a Pre&ndash;Post Survey with District Health Officers in Ghana. J Multidiscip Healthc. 2022 Jul 12;15:1457–68.

12.	Adaletey DL. Leveraging on Cloud Technology for Reporting Maternal and Child Health Services at the Community Level in Ghana. J Health Inform Afr [Internet]. 2017 [cited 2025 May 19];4(2). Available from: https://www.jhia-online.org/index.php/jhia/article/view/146

13.	African Health Initiative Partnership Collaborative for Data Use for Decision Making. Barriers and Facilitators to Data Use for Decision Making: The Experience of the African Health Initiative Partnerships in Ethiopia, Ghana, and Mozambique. Glob Health Sci Pract [Internet]. 2022 Sep 15 [cited 2025 May 19];10(Supplement 1). Available from: https://www.ghspjournal.org/content/10/Supplement_1/e2100666

14.	Country Health Information Systems and Data Use. CHISU Annual Report. Country Health Information Systems and Data Use (CHISU); 2023.

15.	Fähnrich C, Denecke K, Adeoye OO, Benzler J, Claus H, Kirchner G, et al. Surveillance and Outbreak Response Management System (SORMAS) to support the control of the Ebola virus disease outbreak in West Africa. Eurosurveillance. 2015 Mar 26;20(12):21071.

16.	Kaburi BB, Wyss K, Kenu E, Asiedu-Bekoe F, Hauri AM, Laryea DO, et al. Facilitators and Barriers in the Implementation of a Digital Surveillance and Outbreak Response System in Ghana Before and During the COVID-19 Pandemic: Qualitative Analysis of Stakeholder Interviews. JMIR Form Res. 2023 Oct 20;7(1):e45715.

17.	Ampomah S. Vice President launches National E-Health Project to transform Ghana’s healthcare system [Internet]. Ministry Of Health Republic of Ghana. 2024 [cited 2025 May 29]. Available from: https://www.moh.gov.gh/vice-president-launches-national-e-health-project-to-transform-ghanas-healthcare-system/

18.	Agyemang E, Esia-Donkoh K, Adu-Gyamfi AB, Douri JB, Adoma PO, Achampong EK. Assessing the efficient use of the lightwave health information management system for health service delivery in Ghana. BMJ Health Care Inform [Internet]. 2023 Aug 16 [cited 2025 May 19];30(1). Available from: https://informatics.bmj.com/content/30/1/e100769

19.	Amoateng CNA, Achampong EK. Impact of the Lightwave Health Information Management Software on the Dimensions of Quality of Healthcare Data. Healthc Inform Res. 2024 Jan 31;30(1):35–41.

20.	Bawumia launches National E-Health Project to transform Ghana’s healthcare system [Internet]. GhanaWeb. 2024 [cited 2025 May 29]. Available from: https://www.ghanaweb.com/GhanaHomePage/NewsArchive/Bawumia-launches-National-E-Health-Project-to-transform-Ghana-s-healthcare-system-1957949
