## Supplementary material for "The use of data and analytics for health sector priority-setting in Ghana: a qualitative study": S2 Text

**S2 Text: Interview guides**

### Interview guide for academics

[Interviewer: Now, I will define a key term to be used throughout the interview and then we can begin the interview.]

Heath sector priority-setting is the process which aims to “select among different options for addressing the most important health needs…., in the best way, given limited resources” (Schmets et al., 2016).

A. Health governance as it relates to priority-setting.

1. As an academic, what has your role been, direct or indirect, in the process of priority-setting?
2. Which institutions or individuals have you engaged with or supported for health priority-setting in Ghana?
3. Please describe the type of engagement with these institutions or individuals.
4. Please describe the role of other actors you know about in health priority-setting and how they interact. [*Probe for interactions with relation to power, funding and politics*]

 [Interviewer: I will now move on to ask about details of the priority-setting process and approaches used.]

B. Data and analytics

[Interviewer: I will now define another term, and that is “analytics”. Here we are defining analytics as the interpretation of data towards obtaining meaningful information for decision making.]

1. What types of data does your institution provide, if any, towards health priority-setting?
2. What kind of analytic support does your institution provide, if any, towards health priority-setting?
3. Which individuals or institutions do you coordinate with in providing data or analytic capacity towards health priority-setting? This may include institutions from whom data or analytic support is sourced and institutions to which the output of data analysis is reported.
4. Which other organizations do you know about that are involved in providing data for health priority-setting?
5. What types of data do they provide?
6. Which other organizations are involved in providing analytic support for health priority-setting?
7. What types of analytic support do they provide?
8. In what ways do you think the collection and analysis of data for health priority-setting can be improved?

 [Interviewer: I will now move on to ask about support initiatives for priority-setting.]

Here, we are defining support initiatives as entities or projects that provide funding, technical, administrative or other assistance at any step within the priority-setting process.

C. Support initiatives.

[Preamble: There has been local and external support from some organizations to support priority-setting in Ghana. An example is the International Decision Support Initiative (iDSI) which supported Ghana in improving its Health Technology Assessment use.]

1. In your roles on priority-setting, what support initiatives have you worked with?
2. What kind of support has been offered?

D. Challenges and solutions.
1. What challenges have you identified within the priority-setting process in Ghana?
2. (*note*: *If data and analytic gaps were not mentioned in response to preceding question, ask this question*) Are there any specific challenges related to data or analytic capacity?  [*Probe further*: on challenges related to analytic capacity]
3. What are the challenges in communicating analyzed data for decision making?
4. How much do you think data availability influences the timeline for decision making? [*Probe further*: are there examples you can share?]
5. What possible solutions are there to the challenges you have identified?

E. Other.
1. Are there questions I may have missed which might help us obtain more information on health-sector priority-setting in Ghana?
2. Can you please share any additional comments you may have about the priority-setting process?

REFERENCES

Schmets, G., Rajan, D., Kadandale, S. (Eds.), 2016. Strategizing national health in the 21st century: a handbook. Geneva.

Birungi, H., Nyarko, P., Askew, I., Ajayi, A., Addico, G., Addai, E., Jehu-Appiah, C., 2006. Priority setting for reproductive health at the district level in the context of health sector reforms in Ghana. Population Council. https://doi.org/10.31899/rh4.1145

### Interview guide for Ministry of Health and Ghana Health Service directors and program directors

[Interviewer: Now, I will define a key term to be used throughout the interview and then we can begin the interview.]

Heath sector priority-setting is the process which aims to “select among different options for addressing the most important health needs…., in the best way, given limited resources” (Schmets et al., 2016).

A. Health governance as it relates to priority-setting.

1. What has your role been, direct or indirect, in the process of priority-setting?
2. In what ways does your division support national health priority-setting?
3. Which institutions or individuals does your division engage with for health priority-setting in Ghana?
4. Please describe the type of engagement with these institutions or individuals.
5. Please describe the role of other actors you know about in health priority-setting and how they interact. [*Probe for interactions with relation to power, funding and politics*]

 [Interviewer: I will now move on to ask about details of the priority-setting process and approaches used.]

B. Data and analytics

[Interviewer: I will now define another term, and that is “analytics”. Here we are defining analytics as the interpretation of data towards obtaining meaningful information for decision making.]

1. What types of data does your division provide towards health priority-setting?
2. What kind of analytic support does your division provide towards health priority-setting?
3. Which individuals or institutions does your division coordinate with in providing data or analytic capacity towards health priority-setting? This may include institutions from whom data or analytic support is sourced and institutions to which the output of data is reported.
4. Which other organizations do you know about that are involved in providing data for health priority-setting?
5. What types of data do they provide?
6. Which other organizations are involved in providing analytic support for health priority-setting?
7. What types of analytic support do they provide?
8. In what ways do you think the collection and analysis of data for health priority-setting can be improved?
9. How has the work of your division changed over the years in relation to priority-setting?
10. [If interviewee works for the Ghana Health Service (GHS) or the Ministry of Health (MOH), ask the following questions.]
    a. Can you speak about the differences in roles or partnerships between the research and development divisions of the GHS and the MOH as they relate to priority-setting?
    b. How does the RDD at GHS interface with the PPME in priority-setting?

 [Interviewer: I will now move on to ask about support initiatives for priority-setting.]

Here, we are defining support initiatives as entities or projects that provide funding, technical, administrative or other assistance at any step within the priority-setting process.

C. Support initiatives.

[Preamble: There has been local and external support from some organizations to support priority-setting in Ghana. An example is the International Decision Support Initiative (iDSI) which supported Ghana in improving its Health Technology Assessment use.]

1. In your roles on priority-setting, what support initiatives have you worked with?
2. Which of these are local and which are international?
3. [If academia has not been mentioned] What is the role of individuals in academia in supporting the work of your division?

D. Challenges and solutions.
A. What challenges have you identified within the priority-setting process in Ghana?
B. (*note*: *If data and analytic gaps were not mentioned in response to preceding question, ask this question*) Are there any specific challenges related to data or analytic capacity?  [*Probe further*: on challenges related to analytic capacity]
C. What are the challenges in communicating analyzed data for decision making?
D. How much do you think data availability influences the timeline for decision making? [*Probe further*: are there examples you can share?]
E. What possible solutions are there to the challenges you have identified?

E. Other.
1. Are there questions I may have missed which might help us obtain more information on health-sector priority-setting in Ghana?
2. Can you please share any additional comments you may have about the priority-setting process?

REFERENCES

Schmets, G., Rajan, D., Kadandale, S. (Eds.), 2016. Strategizing national health in the 21st century: a handbook. Geneva.

Birungi, H., Nyarko, P., Askew, I., Ajayi, A., Addico, G., Addai, E., Jehu-Appiah, C., 2006. Priority setting for reproductive health at the district level in the context of health sector reforms in Ghana. Population Council. https://doi.org/10.31899/rh4.1145

### Interview guide for development partner representatives

[Interviewer: Now, I will define a key term to be used throughout the interview and then we can begin the interview.]

Heath sector priority-setting is the process which aims to “select among different options for addressing the most important health needs…., in the best way, given limited resources” (Schmets et al., 2016).

For this interview, we would focus on national level health priority-setting in Ghana.

A. Health governance as it relates to priority-setting.

1. What has your role been, direct or indirect, in the process of priority-setting?
2. In what ways does your organization support national health priority-setting?
3. What institutions or individuals does your organization engage with for health priority-setting in Ghana?
4. Which institutions or individuals do your organization engage with for health priority-setting in Ghana?
5. Please describe the type of engagement with these institutions or individuals.
6. Please describe the role of other actors you know about in health priority-setting and how they interact. [*Probe for interactions with relation to power, funding and politics*]

 [Interviewer: I will now move on to ask about details of the priority-setting process and approaches used.]

B. Data and analytics

[Interviewer: I will now define another term, and that is “analytics”. Here we are defining analytics as the interpretation of data towards obtaining meaningful information for decision making.]

1. What types of data does your organization provide, if any, towards health priority-setting?
2. What kind of analytic support does your organization provide, if any, towards health priority-setting?
3. Which individuals or institutions does your organization coordinate with in providing data or analytic capacity towards health priority-setting? This may include institutions from whom data or analytic support is sourced and institutions to which the output of data is reported.
4. Which other organizations do you know about that are involved in providing data for health priority-setting?
5. What types of data do they provide?
6. Which other organizations are involved in providing analytic support for health priority-setting?
7. What types of analytic support do they provide?
8. In what ways do you think the collection and analysis of data for health priority-setting can be improved?
9. How has the work of your organization changed over the years in relation to priority-setting in Ghana?
10. In the use of data and analytics for priority-setting, how does your organization manage tensions between its priorities and national priorities?

Involvement of individual health programs and other sectors.

1. In what ways does your organization interact with program or disease-specific planning offices such as the malaria, HIV and TB control programs for priority-setting?

 [Interviewer: I will now move on to ask about support initiatives for priority-setting.]

Here, we are defining support initiatives as entities or projects that provide funding, technical, administrative or other assistance at any step within the priority-setting process.

C. Support initiatives.

[Preamble: There has been local and external support from some organizations to support priority-setting in Ghana. An example is the International Decision Support Initiative (iDSI) which supported Ghana in improving its Health Technology Assessment use.]

1. Has your organization worked with or developed any support initiatives for priority-setting in Ghana?
2. What types of support initiatives are these?
3. Do any of these involve the capacity for data or analytics?
4. Which of these are local and which are international?
5. [If academia has not been mentioned] To what extent does your organization interact with academia, both local and international, for priority-setting?

D. Challenges and solutions.
1. What strengths have you identified within the priority-setting process in Ghana?
2. What strengths have you identified in Ghana’s use of data and analytics in health priority-setting?
3. What challenges have you identified within the priority-setting process in Ghana?
4. What challenges have you identified in Ghana’s use of data and analytics in health priority-setting?
5. (*note*: *If data and analytic gaps were not mentioned in response to preceding question, ask this question*) Are there any specific challenges related to data or analytic capacity?  [*Probe further*: on challenges related to analytic capacity]
6. How much do you think data availability influences the timeline for decision making? [*Probe further*: are there examples you can share?]
7. What possible solutions are there to the challenges you have identified?
E. Other.
1. Are there questions I may have missed which might help us obtain more information on health-sector priority-setting in Ghana?
2. Can you please share any additional comments you may have about the priority-setting process?

REFERENCES

Schmets, G., Rajan, D., Kadandale, S. (Eds.), 2016. Strategizing national health in the 21st century: a handbook. Geneva.

Birungi, H., Nyarko, P., Askew, I., Ajayi, A., Addico, G., Addai, E., Jehu-Appiah, C., 2006. Priority setting for reproductive health at the district level in the context of health sector reforms in Ghana. Population Council. https://doi.org/10.31899/rh4.1145
