## Supplementary material for "The use of data and analytics for health sector priority-setting in Ghana: a qualitative study": S3 Text

**S3 Text: Interview notes**

### Interview 1

*PATH’s engagement with the health sector*

- PATH conducts landscaping to identify the gaps which exist for a particular issue and then within its own capacity, supports the MoH in addressing the issue.
- This support should be in line with the considerations of the MoH; that is, whatever PATH is doing must be relevant to the MoH.

*How long PATH has been working with in the Ghana Health space?*

- PATH has been working with Ghana since 2002 informally but was registered in 2009.

*How are tensions managed with respect to priorities?*

- Tension never exists between PATH and GHS or MoH, because as an NGO, PATH works in line with national priorities.

*Divisions under GHS that PATH has engaged with.*

- PATH engages with GHS frequently at the national level as PATH does not have staff at district and regional levels.
  - GHS divisions regularly invite the relevant PATH divisions to their divisional meeting.
- Work is in the form of building capacity within GHS to implement PATH activities.
- PATH implements and evaluates initiatives alongside government health agencies.
- PATH engages all GHS departments within their scope. Departments ever engaged include Family and Health Division, Nutrition division and Health Promotion Division, Non-communicable Disease (NCD) program and Research division.
- In PATH’s work, an analysis is conducted to understand the gap of a problem before offering support to the MoH within PATH’s capacities.

*Role of PATH in priority setting*

- *PATH participates in health summits:* The health summit summarises all the happenings in the agencies.
- Before the summit, priorities are already set through the existing health policies and the programme of work which is usually 5 years e.g. salient issues identified from the districts through the regions go to the health summit. The importance of issues is typically assessed using the burden of disease, and this assessment is done with funding and bilateral agencies.
- The final decision is a complex situation. It is done by scoring and the highest score is set as a priority. Global priorities as well as resource availability are also taken into consideration.
- Partner donors pledge their support.
- PATH however influences priorities at the regional and the national level but not at the Health Summit.
- There are other organisations that work with the GHS and MoH.

*Role of PATH in improving capacity for data and analytics*

- PATH has supported to improve data capture tools. For example, PATH experts work with the MoH to add indicators to the government’s data system. Also, PATH experts help to identify gaps and support the ministry to strengthen work in these areas.

*Strengths of Ghana’s health priority setting system*

- Use of a bottom-up approach: priorities are set at district, regional and national levels.

*Weaknesses of Ghana’s health priority setting system*

- Political influences on the priority-setting process.

### Interview 2

*Type of data that comes to the PPME*

- The aggregate data comes from the District Health Information Management System (DHIMS2) which captures data from all facilities under the Ghana Health Service (GHS). Data from the teaching hospitals are on a different system.
- The DHIMS2 data includes both public and private facilities data.
- However, not all private facilities are currently reporting their data.
- Health Facilities Regulatory Agency (HEFRA) mandates all facilities to report into the DHIMS.
- The DHIMS data comes from routine activities.
- Administrative data collected from the health facilities include, morbidity, mortality, maternal, mortal audits, post-natal, institutional neonatal births, immunization (BCG, Penta 3, etc.), top 10 or 20 causes of morbidity and admissions which are the key routine data.
- Also, epidemiological data, which include incidence, prevalence, mortality and cause of mortality are included.

*Data analysis objectives*

- The data is analyzed based on the objectives of the meeting the PPME is presenting at. Analysis is also based on the objectives of the request received from GHS or MoH directors who require data aggregates for specific meetings. For example, when the Director General of the GHS is preparing to attend a meeting, the data is analyzed by the PPME based on the requirements of the meeting he is attending.
- Sometimes, PPME adds other indicators to the DHIMS depending on needs or identified gaps. For example, routine data did not include some indicators in the past but due to the Primary Health Care program, some have been added recently.

*What PPME does to the data collected*

- Data is first validated to check accuracy of the data. Facilities are contacted for verification, if need be, through the health directorates at the various levels.
- Analysis is then performed based on requests, e.g., trend analysis, spot map using GIS component of DHIMS 2, graphs or pie charts.
- Analysis is also performed to assess the timeliness and completeness of the data.
- Examples of groups of aggregated data to inform priorities: Disease burden data, health service utilization data and demographics.

*Agencies that request for data*

- Requests come from various institutions and agencies.
- They ask for the specific data they need based on their objectives.
- Requests are also made by universities for their research work.
- Other divisions of the GHS also request data.

*Interactions with external and internal support agencies*

- Global fund, World Bank, World Health Organization (WHO), United Nations International Children's Emergency Fund (UNICEF), Korea International Cooperation Agency (KOICA) provide support with the following.
  a. Data management.
  b. Capacity building of the staff in data analytics.
  c. Policy development in relation to data management.
  d. Logistic support such as provision of tablets, laptops internet connectivity, fund server space.
  e. Organization of routine meetings
  f. Health system strengthening.
  g. Presence at management review meetings.
  h. Supportive supervision on the field.
  i. App development (app used for collecting data on the field).
- The PPME is usually invited to GAVI and UNICEF planning meetings.

*Interaction with Ministry of Health (MoH) Research Statistics and Information Directorate*

- Ministry has direct access to the DHIMS to generate information from there.
- They contact the GHS-PPME when they have difficulty in accessing an aspect of the DHIMS.
- The GHS-PPME submit reports to them when MoH requests for it.
- GHS-PPME and MoH support each other in certain meetings.
- The MoH shares priorities and targets with the GHS-PPME.

*Agencies outside the Health Service that receive data*

- Ghana Statistical Service
- Parliament of Ghana
- Ministry of Finance through the MoH.
  - The MoH submits two reports quarterly to the Ministry of Finance: financial report and non-financial report.
  - The non-financial report consists of the activities the ministry is doing within the quarter and what has been done so far. In these reports, there is the budget implementation report, which includes a section of data from the PPME with activities and indicators.

*Is PPME invited to the Health Sector Medium-Term Planning Meeting (HSMTP)?*

- All agencies under the MoH are included in setting the document on health priorities.
- All agencies are expected to report at these meetings their program of work. The HSMTP final document is an overall plan of work.
- In this document, agencies are attached to various programs.

*Is the PPME office specifically invited to the HSMTP?*

- Yes, since PPME is the hub that generates all GHS data documents.
- Other agencies are called to join the PPME if need be.
- The PPME is the voice for the GHS.

*Factors that play when directors are deciding what the GHS should prioritize*

- PPME presents at meeting based on the objectives of the meetings. This presentation is a report on relevant (specific to objectives) indicators.
- Any division that also presents at the meeting presents on the indicators in line with the objectives.
- Resources also play a role in the indicators that have to be presented on.
  - As an illustration: Out of the funds allocated to the health sector, close to 70% goes to personal emoluments and only about 30% is left for activities within the ministry.
  - Then the over 23 agencies have to share the 30% across for their activities.
- The development partners also have their priorities and are interesting in funding their priorities. (For example: USAID prioritizes health system strengthening, monitoring and evaluation while the World Bank prioritizes programs such as HIV, TB, etc.)
  - So, depending on who is bringing the money and where their interest is, a chunk of the money is directed to that.
- Though the ministry can sometimes allocate some amount to other priorities, it is very limited due to the limitations in overall government funding.

*How capacity is built in relation to data analytics currently in the PPME*

- Depending on the level of capacity desired to be built, the relevant agency sets the “how”.
- At the national level, PPME determines the content of training.
- This kind of capacity building occurs about twice a year, but actual frequency is dependent on the feedback from routine supervision and funding.
- Specific capacity-building trainings that have been organized:
- Data management training
- Training on Standard Operation Procedure (SOP)
- Update and support supervision comes with refresher trainings.

Capacity building

1. *Digital health training*

- Currently, the GHS is operating the digital health which comes with the ICT department.
- The department is training officers at various levels using the e-learning platform.
- The e- way of training has been operational for 6 years with over 14,000 workers subscribed.
- The digital health department was formally a unit under the office of the Director General of the GHS but is currently operating as an established department under the PPME.
- Involves training in digital tools in Surveillance, Outbreak Response Management and Analysis System (SORMAS).
- Training in capacity to provide support to SORMAS.
- Training in diagnostics and descriptive analysis.
- Currently, the effort is to move towards training in predictive and prescriptive analysis.

2. *Trainings on DHIMS*

- Before any form is added to the DHIMS, PPME pilots the form in selected facilities and training is done before utilization.
- Assessment of the system is done twice a year to test the gap and improve on it and after staff are trained on the data quality process.
- Capacity is also built on using GIS in the DHIMS. The health facilities are identified with their coordinates.

*Interaction with academia*

- There are partnerships with academia in conducting research.
- There is a partnership with University of Ghana School of Public Health.
- There are collaborations with institutions of research.
- Also, with Kwame Nkrumah University of Science and Technology as consultant to provide technical assistance through the UNICEF on the Water, Sanitation and Hygiene (WASH) Infection Prevention and Control (IPC) program.

*Outcome of academia interactions*

- Publications
- Policies for health promotion, etc.
- Policy dialogues, policy briefs with the KNUST program.

*Challenges*

- There is need for continuous support for the space on the server.
- Funding for internet connectivity.
- Currently operating both the paper based and electronic system and this generates a volume of work for GHS staff. Funds are needed to completely move to the electronic system.
- Unable to do routine research for the system due to lack of funds.
- Lack of data analytic capacity for health information officers to generate visuals from data to better understand current trends and inform decisions.
- Data coverage is not a problem but the visualization of the data to increase usability is the problem. Support is needed to build capacity of staff to enhance data accuracy and visualization.

*What is done currently about challenges.*

- Donor partners are asked to support fund space on the server.
- Currently, health facilities are funding that but they still have issues because the National Health Insurance Scheme (NHIS) Funds are not paid on time.

*Other notes:*

- Planning and budgeting are very important in priority setting; hence it should be considered in this project.
- There are health planners and health information officers at the various health care levels who help manage the data at those levels. As there are no dedicated PPME offices/officials at the sub-national level, the directors at these levels act as the PPME at their levels by helping to translate data policies from the national level to the sub-national levels.

### Interview 3

*Overview of Research and Development Division of the Ghana Health Service*

- Research and Development Division (RDD) is one of the 11 Ghana Health Service (GHS) divisions.
- RDD has 3 branches.
  - Research Coordination
  - Documentation, dissemination and advocacy
  - Ethics and research management
- Role of RDD: Generate data/information to inform policy and ethics.
- RDD oversees the three health research centers in Ghana (Dodowa Health Research Center, Navrongo Health Research Center and Kintampo Health Research Center).

*Role in Priority-Setting*

- RDD generates research data to support decisions.
- There are platforms such as the Demographic and Health Survey (DHS) that are used to generate such information.
- Commercial agencies also seek research data from RDD. These do not always aim at supporting the national agenda. They may support the international agenda. If platforms generated information.
- The main evidence is presented through the documentation, and dissemination and advocacy unit to the Policy, Planning, Monitoring and Evaluation (PPME) division and MoH; and then used to inform policy briefs.

*Main audience*

- The division has three departments.
- In ideal circumstances, RDD’s work should be as follows.
  - Generates data from research centers.
  - Documentation branch documents the data.
  - Research dissemination branch disseminates the research.
  - Advocacy branch *synthesizes research to policy brief.*
  - Policy briefs is forwarded to the PPME.
  - PPME develops policy and coordinates implementation.
  - PPME is supposed to be a policy implementation division, but this is not the case.
- The research division would have done greater good if positioned at the MoH, given that the MoH is the policy making body.

*Development and Delivery of National Health Plan*

- The PPME coordinates all activities from all the divisions of the GHS. All divisional directors meet at the National level to present their reports. This meeting is chaired by the Director of the Ghana Health Service.
- Sometimes priorities are set based on reports from the various units. The Director General of the GHS sets the agenda for the meeting.
- Mostly the Minister of Health and Government of Ghana are not able to deliver the National Health Plan due to limited resources.

*At what stage is the RDD involved in priority setting?*

- Ideally, RDD should be providing the data for priority setting. However, at MoH meetings, other agencies (aside the GHS, such as Teaching hospitals, child unit) submit their own findings because data does not sit with one agency.
- How priorities are set from the GHS to national level:
  - RDD findings feed into the general GHS report at the national level. For meetings at the national level, which are chaired by the minister of health, the director of research might not even be needed given that their reports will be represented by the PPME or Director of the GHS. The Director General’s office is coordinated by the PPME.
  - PPME staff often attend the MoH’s meeting on behalf of the GHS with the Director General.
  - The PPME acts as a secretariat to the GHS and collates all the reports from the various 11 divisions.
  - The Director General then decides which reports or health areas to present at meetings with the MoH.
    - Priorities are generally dependent on the burden of the problem, defined by the data.
    - Routine surveillance data is managed by the PPME.
  - The MoH will also set their priorities (based on reports from the GHS, other agencies) and then the minister presents to the government.
  - Finally, the government performs the final priority setting.
  - Funding also determines what the priorities will be.
    - Priorities are often set to align with a funder’s agenda.
- Within the RDD, priority setting is conducted based on annual reviews at the national level (RDD headquarters in Accra, including its 3 branches) and at the research centers.

*Relationship within RDD GHS and Research Statistics and Information Management (RSIM) at the MoH?*

- Regulation: RSIM MoH should be coordinating/regulating all the health research in the country; but, rather they conduct their own research
- Why does RSIM MoH conduct their own research?
  - Research is an avenue for attracting funding.

*Challenges with analytical capacity*

- Much is not done in the area of advanced analytics on the data because most audience will be unable to understand the results.
- There are a lot of questions to be asked for policies to be made, but there is over reliance on little data. In-depth analysis is not appreciated.
- There is also little expertise in that area. We lack critical analytics. Research and analysis are not requirements to be a General Director.
- The GHS does not have a lot of qualified statisticians which is quite difficult. i.e. limited critical analytic experts in the system.
- Universities are the best places to build analytic capacity. Currently, it is difficult to attract PhD holders to the GHS and there is high staff turnover.
- There is no motivation for staff to advance their analytical skills, as there is no demand for advanced analytics on the GHS data.
- MoH does not have a lot of technical people.

*How can analytic capacity be developed?*

- Capacity should be built in the Universities so that researchers can partner with health agencies like the GHS in the analysis of the data they generate.
- Analytic skills are needed at all agencies, including the teaching hospitals
- To increase analytical capacity in the GHS,
  - there should be partnerships between the universities and the GHS.
  - there must be a common platform to analyse and synthesise data.
  - possessing analytical skills must be a requirement to be recruited to specific positions that require those skills.
  - there must be incentives like appointments and promotion based on analytic capacity so people will want to do analytics.
  - there must be training in critical thinking rather than only interpreting basic proportions and frequencies.
  - At higher levels of governance, individuals must possess capacity for analytics.

*Challenges with data and analytics for priority-setting in Ghana*

- People lack interest in analytics because people do not value analytics in their work to inform policies.
- Priorities are also politically driven and not really evidence based. Priorities are often set where weight is placed, either politically, economically or externally.
- Local funding is a challenge. Therefore, most of the priorities set are influenced by external funders such as World Health Organization (WHO), JICA, KOICA, etc. Example: Vitamin A, bed nets, artesunate for malaria
- Funding and “big players” drive priority setting.
- When there is money backing evidence, it is moved to policy.
- “Research findings are often left on the shelves. It informs policies when it’s an emergency or when it has a lot of funding to push it to the PPME”.
- Funding is needed for training and other things at PPME. Some policies are informed only by evidence from other countries and then implemented in Ghana through WHO, as it is a major funder.
- When findings come from other WHO member states and policies are made, Ghana like other low-resourced settings is forced to accept them as policies.

### Interview 4

*Stages in priority setting*

- Priorities are set based on existing national health policies, the health sector medium term development plan (HSMTDP) and global priorities such as the Universal Health Coverage.
- The HSMTP is developed based on the National Health Policy and UHC roadmap.

*Role of data and analytics in priority setting*

- Routine data is mainly used for assessment of performance and health system targeting
- Priorities may also be set based on findings from the literature.

*Role of MoH in priority setting*

- Key source of data has been District Health Information System (DHIMS), which is managed by the GHS.
- However, this data source does not include data from teaching hospitals, as they manage their own data and are hesitant to reporting to the GHS DHIMS. Military hospitals also fall within this category. Altogether, there are about 400 facilities not reporting to the DHIMS.
- Therefore, the ministry is currently working on an information system that will merge data from various agencies such as the teaching hospitals and other quasi-institutions.
- The priority setting process:
  - Data from the GHS and other agencies are analyzed and presented to the MoH.
  - Depending on the identified indicators, an ad hoc committee is formed with the Policy and Planning, Monitoring and Evaluation director as chair.
  - The output of this committee is an assessment of the health situation.
  - The assessment contributes to the “Aide Memoir”, a list of priorities developed by the government in consultation with funders at the national health summit.

*Challenges with data and analytic capacity for HSPS*

- The effectiveness of the indicators in the DHIMS data needs to be reassessed. For example, there are few or no indicators on the patient experience.
  - There are many indicators, but many are inadequate.
- There is a gap in merging data from various agencies.
- Need for local expertise in big data analytics and presentation.
  - We typically rely on counts and averages; we are not measuring relationships.
  - Technical capacity is mostly external.
- Unavailability of program data.
- Research input by health partners is also unaccounted for.
- Policies are made too fast with little dependence on data.
- There is no problem with data generation, the main issues are bringing them together, assessing, and translating them into policy.

*How can data and analytic capacity be improved?*

- Developing a national health database which brings data from multiple sources.
- Improving local technical capacity
  - Also, capacity in visualizing data
- A capacity needs assessment is needed.
- Developing analytic capacity among decision makers

*Support for priority setting*

- The USAID provides the following.
  - financial help in the form of equipment acquisition.
  - technical help in the form of consultancy.

*Additional thoughts for the project*

- Important to clarify the level at which we are speaking about setting priorities.
- We also want to think about how data helps with the implementation of set priorities.

### Interview 5

*Overview of Priority Setting in Ghana*

- Note that the process to be explained may not be the current practice.
  - Information is based on interviewee’s experience when they were working with the GHS.
- The planning process starts with the five-year health sector strategic plan.
- Annual work plan is developed from the strategic plan.
  - Annual plan is developed in consultation with various stakeholders.
  - Key lead health sector stakeholders meet every month to deliberate based on prevailing health trends or news.
    - It is a very dynamic policy implementation forum for prioritisation.
    - Priorities are influenced by the global agenda, current news and secured funding.
    - This happens monthly.
  - Each year's plan will have various thematic areas based on the priorities of the health sector.
- The priorities emanate from various actors, e.g. the health sector working group, and inputs are collected from all agencies and partners
- Priorities must be linked to available finances, which means that the priorities of partners must be taken into consideration i.e. USAID, World Bank etc
  - For example, in the past, a funder had five regions in Ghana as its priority regions, based on poverty levels in those regions. Hence, the funder supported work only in those regions during that period.
- Priority setting is also determined by global health agenda and associated international programmes. Countries do not want to be isolated from the global agenda and hence set priorities to align to these.
- Over the years, the Ministry of Health became very weak in setting health priorities. The following are causes of this.
  - Priorities were set by the partners and other agencies.
    - For example, agenda 2030 (Sustainable Development Goals [SDG]), advocated for Universal health Coverage. Once this global agenda was set, it meant the annual plan had to be revised to incorporate the SDGs.
  - Political interference was present. Although the government’s manifesto might be different from the prevailing health agenda, the health agenda will often be revised to incorporate manifesto promises, even if that did not reflect prevailing needs.
    - E.g. In 2018, during the annual health summit, the Vice president mentioned the use of drones to deliver vaccines which was not a priority then. Policies then needed to be developed in retrospect to accommodate the Vice President’s comments.

*Annual Performance Review (APR) and priority-setting*

- The APR is an implementation and policy platform.
- A 2-year trend is done to assess the performance indicators against priorities.
  - Results of this assessment tell how the health sector is performing.
- This assessment happens at the district, regional, and national levels. District data is collated at the regional level, and regional data is collated at the national level.
- The APR reviews are collated and present at the annual health summit.

*Annual health Summit and HSPS*

- It is held once a year by all health sector players including the partners and is chaired by the Minister of Health.
- Priorities are set during this summit.
- Summit at which “national picture” of health is presented.
- On the 4^th^ day of the summit, participation is limited to the minister, agencies and partners who in a meeting called the “Partners Forum”, to deliberate on priorities. Technocrats (GHS and MoH staff at the national level) may also be present. The outcome of the forum is the Aide Memoire, a signed document on agreed priorities and funding commitments.
  - In the forum, partners make commitments based on their own priorities.
- Funding commitments in the memoire help to give a fair idea of how much money is coming to the ministry.

*What is used for priority setting*

Global agenda

Routine surveillance

Assessments (Performance Reviews)

- Aside the APRs, PPME also conducts a quarterly assessment of the health service based on DHIMS.
  - There are alert systems within DHIMS.

*Priorities and Finance*

- Before the health sector reform, the MoH managed its own finances.
- After the health sector reform, health finances are managed and released by the Ministry of Finance (MoF). GHS has drawing rights but need approval from MoF.
  - Regions and districts are required to present a budget to the MoF.
  - PPME liaise with regions and districts to prepare the budget and priorities.
  - Sub-district level collate budgets to the regional level and then regional level collates to the national level.
  - GHS defends its budget at a hearing.
  - MOF exercises discretion in disbursing funds.
  - All requested funds are seldom provided because government usually prioritises salaries which make up about 94% of annual health budgets, the rest goes to infrastructure.
- When partners provide funding, the Ministry of Finance could reprioritise where to use the funds and only needs to write to the partner to inform them about the change in purpose of the funds.
  - Do partners agree to this change? Yes, mostly.
  - It is worth noting that the minister of finance is powerful and can determine what to prioritise.
  - For example, WHO funds for the Maternal and Child Health and Nutrition Project were repurposed to fund COVID-19 control efforts.
- There have been a lot of efforts at prioritisation but there is no matching finance for implementation.
- At the national level, there is prioritisation but there are specific areas of focus. Sometimes, these are influenced by available resources. The situation is complex within the national system.
- When priorities align with those of funders, the advantage is that resources will be available to execute priority-related activities.
- You can prioritise but if you do not have resources, it will only be on paper.
- Priorities without funding is not good.

In Ghana, dependencies on donors have been very strong.

*National Health Insurance Scheme*

- The current Health Insurance is purely used for preventive care. To bridge the gap in the current model, it should include promotion, prevention as well as curative.

*Data and analytic capacity for priority-setting*

- In the past, there was very little effort to start looking critically at data at the point of care; for example, to know that increased incidence of a particular disease may indicate an outbreak. The data was simply collected and packaged to the DHIMS.
- In 2016/17, community data collecting tools (tablets) were introduced in the communities to collect data and generate graphs. This introduced a team approach to data analysis.

*Challenges*

- Leadership
- Inadequate technical capacity on human resource
- Political interference
- Disconnect between the local and national levels.

What gets prioritized at the national level is not implemented at the local level.

*Way forward*

- A strong health sector leadership will help in priority-setting
- We need to stop seeing health as a business commodity and see it as a social intervention.
- There is a need for a strong PPME at the ministry level to be able to perform analysis and develop a priority implementation plan.

*Overall point*

- Priorities may be identified based on these data, but the resources may not exist to implement this.
- Overall point: Prioritization is driven by available resources.

### Interview 6

*Relevance of research (done by UHAS researchers) to policy*

- Personnel from the GHS are involved in the research process and become co-authors of papers. They are invited to dissemination programmes.
- The centre for Health Policy and Implementation Research (World Health Organization) is set up to train young faculty in implementation research.
  - To increase relevance of research to policy

*Strategies to increase relevance of research to policy issues*

- Engaging with the existing health sector plan of work.
  - Stakeholders will be more willing to engage of research aligns with health sector plan
- Engaging in stakeholder consultation processes prior to beginning one’s research in order to learn about any process of prioritization of issues which researcher may be unaware of. Areas to consider:
  - *Relevance of the issues*
  - *Whether someone has done it before*
  - *Timeliness of the issue*
  - *Financial restrictions*
  - *Interest*
- Selecting the right stakeholders to engage with.
  - Important to not only target the head of the organization but also others in the organization, in case leadership changes.
- Making sure to involve as many people as possible to incorporate diversity of thought.

*Challenges in research for policy*

- People talk about research but may not understand the process. They usually focus on their comfortable space forgetting other areas e.g. the labs.
- Sometimes, there is lack of monitoring of the research.

*Strengths of the priority Setting process in Ghana*

- The attempt to set priority is in the right direction but implementation is the main issue.
- The health sector has the DHIMS to integrate the data very well and ensure data informs the process and the funding.
- For the effective implementation of Aide Memoire, monitoring and evaluation is very important.

### Interview 7

*Role of WHO Ghana in priority-setting*

- Supports the Ministry of Health (MOH) to generate, synthesize and analyse the data they generate to ensure the monitoring of health quality and SDG attainment.
- Supports the MOH and its allied agencies to perform analytics to inform programs and policies.
- Ensures research ethics, norms and standards of WHO are adhered to.
- Supports data quality
  - WHO supported research studies to go through rigorous quality processes.
  - Quality is not an issue with research data but administrative data like disease burden is the issue. The Ghana Health Service (GHS) has standards and audits that these routine data should go through, but a survey might be needed to ascertain whether data goes through these checks.
  - Most of the times, what has been observed is that the data is accurate with just little acceptable levels of errors.
- Helped to support the country on harmonizing health facilities assessment, STEPS, DHS, subnational unit assessment (so that districts know where to place their priorities)
- Supports the MOH in implementing its policies. The support is given on a day-to-day basis.
  - For example, ensuring 1 billion more people get access to Universal Health Coverage (UHC).
- Strengthening health information systems to support MOH, its agencies and other non-health agencies such as Ghana Statistical Service, Ministry of Gender and Ministry of Education, etc.
  - For example, for the past two years, the WHO has helped the country to generate data for priority setting using the SCORE Assessment.
  - For example, in 2018 SCORE assessment for Ghana was 65%.; i.e., Ghana did not have 35 percent of data needed on monitoring progress towards the Sustainable Development Goals (SDGs).
    - The WHO is helping to bridge that gap.
- Helps with sentinel services and supports the Ghana DHS and sub national units at district levels to determine where to place their priorities in terms of UHC. That is, moving from the data and analytics to what needs to be done based on the analytics results.

*Engagement with the MoH:*

- WHO engages with the MoH to share in their priorities and inform them of the key focus areas of the WHO.
- WHO supports the planning cycle of the Health Sector Medium Development Plan.
- WHO priorities are set based on country data and the Health Sector Development Plan.

*How tension between national priorities and WHO priorities resolved*

- WHO does not operate in a vacuum, the priorities from WHO are generated from country data.
- The WHO Health Center Development plan, SDGs, etc. all align to government priorities.
- There is usually no tension in priorities. The problem has been with the order of priorities; in which case, discussions are held to decide.

*Support WHO provides in terms of data and analytics*

- WHO does not generate and provide data.
- It rather provides tools and technical support to Ghana to be able to use the data the country generates.
- WHO provides technical support for analysis and synthesis to move data to knowledge generation in areas the WHO thinks there is a big gap.
  - WHO provides the expertise and skills needed for using data in developing policies and interventions.
  - Generation is one stage; analytics is another and usage is another. Usage requires the skills and expertise.
- The WHO provides training in using data for policy briefs, info graphs, etc. to help people understand and to improve uptake of data.
- WHO also provides capacity building as well as consultancy services to train managers and other stakeholders from the MoH to analyse the data.
  - The frequency of these trainings depends on needs and requirements for the ministry and is tailored to specific needs.
  - The WHO has organized the following capacity building workshops this year.
    - District health functionality assessment:
    - Harmonized health facility assessment
    - Ghana Demographic and Health Survey training: Training on using access mode and QGIS for accessibility for emergency referrals from Community-Based Health Planning and Services, etc.
    - Health inequality monitoring
      - Training on a health equity assessment tool to help monitor health inequalities to ensure that the amount of knowledge generated from available data tells a story to help people understand the status quo.

*Interaction with disease-specific Programmes*

- Most workshops are typically sector-wide approaches and not disease-specific.
- However, if a disease programme needs special support, WHO offers it.
- All activities of WHO have to be linked to the health sector objectives.

*How WHO interacts with academia*

- One of WHO KPI’s is involving academia in its programs.
  - WHO Ghana has worked with UHAS, Noguchi Memorial Institute for Medical Research, University of Ghana School of Public Health, and Institute for Statistical, Social, and Economic Research (ISSER).

*Strengths in the priority setting process in Ghana*

- Well established cycle and structure for planning and budgeting.
  - However, it is hard to ascertain whether the structure is working.
- It is good that the GHS priority setting process allows a bottom-up approach, with contributions from sub-districts to district to regional to national levels.

*Challenges of the priority setting process in Ghana*

- Inadequate funding for implementation of set priorities
  - This can be a demotivating factor to setting priorities.
- Leadership and governance are a big issue. It is good from the bottom till you get up. There is an issue of apathy at the national level.
- Translation from data to policies:
  - There is a big gap between data analysis and knowledge production, i.e., developing knowledge products such as policy briefs, infographics
  - Ghana is rich in data but capacity in how to use the data is lacking.

### Interview 8

*TB Program background*

- Tuberculosis (TB) is under the Public Health Division, which has these three disease-specific programs:
  - TB
  - Malaria
  - HIV

*Programs and their contribution to the national health sector plan*

- Individually each program has an institutional strategic plan which contributes to the GHS strategic plan.
- GHS plan contributes to the MoH plan.
- MoH sets the overall health agenda.
- The United Nations (UN) Sustainable Development Goal (SDG) 3 is a reference point for the MoH plan.

*Priority-setting within the TB program*

- TB prioritization follows the WHO method of standardization.
  - Patient centered framework prioritization (PCF) is what is used
    - What is the PCF? It involves:
      - Incorporating the views of the patient and patient experiences.
      - Consulting at community, subdistrict and district levels, and speaking to community leaders, including chiefs
- The following groups are also consulted in priority setting.
  - Health workers
  - Ministry of finance, for budgeting purposes
  - Parliamentary caucus
  - Ministry of Gender, Children and Social Protection
    - Why is this ministry relevant?
      - It manages the Livelihood Empowerment Against Poverty (LEAP) program and given that TB is considered a pro-poor disease, the TB program partners with this ministry.
    - National Health Insurance Authority (NHIA)
- In the planning process, the first thing is to invite technical experts, e.g., USAID, WHO, WHO-Afro-Region, KNVC) to offer technical assistance.
  - This assistance provides support in organizing the mid-term review (epidemiological review) and other technical analyses such as patient pathway.

*What data is used for prioritization?*

- Routine surveillance
- Data on high-risk groups: e.g., prison inmates, pregnant women
- Time since diagnosis
- Diagnosing physician

*Tool for data entry*

- Case based data entry called e-Tracker

*Capacity for data and analytics:*

- Policy, Planning, Monitoring and Evaluation (PPME), GHS, manages capacity for Utracker and District Health Information Management System (DHIMS)
- Capacity has been improved at the level of the health facility for use of a TB screening tool for intensified case finding.
- Capacity is being strengthened in modeling TB incidence estimates to improve the burden estimates for the country. This is a partnership with the WHO.
- Capacity improvement needed:
  - Among lab scientists and nurses to improve data surveillance
  - Among referral clinicians to more easily identify a case

*Data and analytics challenges for the TB program*

- Need for better TB burden estimates
  - Actions toward correct incidence estimation

Intensified case finding

Sample transportation

Contact tracing

Focus targeting on hotspots (mining area, prisons, facilities with more cases)

Effect of other diseases, COVID-19 example: TB tests during COVID-19 testing era allowed more TB cases to be detected.

- Resources: main funding for program is drawn from Global Fund alone.
  - Program activities are not funded by government; only personnel salaries are.
  - Program is underfunded by 45%.
  - There are no funds for operational research.
  - Some agencies have directly supported the program in the past. For example:
    - UNICEF began to provide stool testing for TB.
    - IOM provides cross-border TB testing.
- Reporting: Of the 2000 Directly Observed Therapy (DOT) centers for TB, only 425 report their data electronically. This is connected to the financial resources challenge.

*Interactions between disease programs in terms of data and analytics*

- There is sharing of data.
- However, collaboration is challenging because the donors to specific programs define what the agenda should be. If collaboration is not a priority of the agenda, this may not happen.
  - For example, TB/HIV Grant: The TB and HIV programs communicate as stakeholders, but funds may not be shared.

*How data and analytics challenges can be improved*

- Ghana government contributing to program funding.
- Task shifting; allowing health workers to train for and execute specialized tasks as needed. E.g., a nurse learning how to input data so they can cover for the data manager if needed.
- Ability of disease programs to communicate with one another clearly in terms of their data: for example, if one has malaria, we want to know how much of those cases have TB through the DHIMS
- Greater coverage of e-Tracker

*Additional notes*

- Note that Ghana contributes to the TB world report based on empirical data (district, regional).
